# Kidney Failure Prediction: Multicenter External Validation of KFRE Model in Patients with CKD Stages 3-4 in Peru

**DOI:** 10.1101/2023.03.27.23287771

**Authors:** J Bravo-Zúñiga, R Chávez-Gómez, P Soto-Becerra

**Affiliations:** nephrologist at Nephrology Department, Hospital Edgardo Rebagliati Martins, EsSalud, Lima, Perú; associated researcher at Universidad Continental, Huancayo, Perú

**Keywords:** Prognostic model, chronic kidney disease, chronic kidney replacement therapy, Kidney Failure Risk Equation

## Abstract

**Background:** To externally validate the 4-variable Kidney Failure Risk Equation (KFRE) in the Peruvian population for predicting kidney failure at 2 and 5 years.

**Methods:** We included patients from 17 primary care centers from the Health’s Social Security of Peru. Patients older than 18 years, diagnosed with chronic kidney disease (CKD) stage 3a-3b-4 and 3b-4, between January 2013 and December 2017. Patients were followed until they developed kidney failure, died, were lost, or ended the study (December 31, 2019), whichever came first. Performance of the KFRE model was assessed based on discrimination and calibration measures considering the competing risk of death.

**Results:** We included 7519 patients in stages 3a-4 and 2,798 patients in stages 3b-4. KFRE discrimination at 2 and 5 years was high, with Time-Dependent Area Under the Curve (AUC-td) and C-index > 0.8 for all populations. Regarding calibration in-the-large, the Observed-to-Expected (O/E) ratio and the calibration intercept indicated that KFRE underestimates the overall risk at two years and overestimates it at 5-years in all populations.

**Conclusions:** The 4-variable KFRE models have good discrimination but poor calibration in the Peruvian population. The model underestimates the risk of kidney failure in the short term and overestimates it in the long term.

**SIGNIFICANCE STATEMENT:** The Kidney Failure Risk Equation (KFRE) is a widely used prediction model for kidney failure risk assessment in patients with chronic kidney disease (CKD). However, its performance in Latin American populations remains unclear, particularly in primary care settings. This study externally validated the KFRE in Peruvian CKD patients, demonstrating high discrimination but revealing miscalibration that could lead to adverse patient outcomes resulting from over- or under-estimation of risk. These results underscore the need for model updating and further research to optimize the KFRE’s use in clinical practice in Latin America, provide valuable insights for applying the KFRE in Latin American settings, and highlight the importance of continuous evaluation and refinement of prediction models in diverse populations.

## INTRODUCTION

Chronic kidney disease (CKD) is a major global public health challenge, imposing substantial economic burden on healthcare systems and displaying an ever-increasing prevalence [1,2]. With over 10% of the global population affected [2–4], the burden of CKD is particularly pronounced in low- and middle-income countries, where resource constraints and fragmented healthcare systems exacerbate the issue [2,5,6]. In Peru, a Latin American middle-income country, over 2.5 million adults suffer from varying degrees of CKD, with regional prevalence estimates ranging between 16% and 20% [7–10].

Timely referral of CKD patients to nephrologists significantly reduces healthcare costs as the disease progresses towards kidney failure and death. Early referral is crucial in healthcare systems with limited health networks and scarce nephrology specialists [11]. CKD progression risk prediction models assist clinical decision-making by informing nephrologist referrals, providing kidney replacement therapy (KRT) counselling, and aiding in vascular access planning to prevent sudden, unplanned emergency admissions [12,13]. Accurate short-term predictions are particularly important for establishing individualised risk [14–16], while long-term predictions help identify patients requiring primary care for secondary prevention, treatment, and follow-up [14–16].

International guidelines recommend individualised risk prediction models to determine the optimal timing for nephrologist referral and KRT planning [17–20]. However, in Peru, referral recommendations rely mainly on eGFR, albuminuria, or ACR thresholds, either in isolation or combination [21]. These non-individualised criteria may lead to unnecessary referrals of low-risk patients and missed referrals of high-risk patients [17,22].

The Kidney Failure Risk Equation (KFRE) offers an individualised risk prediction for CKD patients’ progression to kidney failure [23] and is recommended by some international guidelines for CKD management [14,18,19,24]. The KFRE has 8-variable and 4-variable versions, with the latter being an appealing option for the Peruvian health system, requiring only a few easily accessible variables. However, predictive errors can vary significantly between populations, emphasising the need for validating their predictive performance in the target population [25–27]. The absence of KFRE external validation studies in Latin America, including Peru, has delayed its adoption in local clinical practice.

This study aims to externally validate the predictive performance of the 4-variable KFRE model in predicting 2 and 5-year risk of kidney failure within a large and heterogeneous cohort of patients presenting with CKD stages 3a, 3b, and 4 in primary care settings in Peru.

## METHODS

### Design, Study Population, and Data Source

We conducted a retrospective cohort study following the Transparent Reporting of a multivariable prediction model for Individual Prognosis Or Diagnosis guidelines (see TRIPOD checklist in **Supplementary Material**) [27,28] to validate the Kidney Failure Risk Equation (KFRE) model for predicting the risk of kidney failure at 2- and 5-year horizons in patients with chronic kidney disease (CKD).

As part of the EsSalud National Kidney Health Plan, the Rebagliati Healthcare Network established an electronic registry of CKD patients treated at its affiliated health facilities from primary care until tertiary level. We obtained demographic and clinical data from the electronic medical records of the EsSalud Hospital Management System and the Chronic Kidney Disease Management Unit (UMERC) of the Kidney Health Surveillance Subsystem (VISARE) computer application [29].

The study population included patients aged 18 years or older with CKD between January 1, 2013, and December 31, 2017, treated in 17 primary care healthcare centers in the Rebagliati Healthcare Network in Lima, Peru. Lima is the capital of Peru and concentrates most insured patients at national level. We included patients with an estimated glomerular filtration rate (eGFR) between 15 and 60 ml/min/1.73m^2^, corresponding to categories 3a, 3b, and 4 of the Kidney Disease Improving Global Outcomes (KDIGO) classification [20], and those who had a recorded quantifiable albumin-to-creatinine ratio (ACR) measurement taken simultaneously with eGFR. The date of ACR measurement was the start point for beginning the follow-up and for predicting the risk of kidney failure using KFRE.

We validated the KFRE models in two populations of interest: (i) a broad population of patients with CKD stages 3a, 3b, and 4 (3a-4), such as originally was validated, and (ii) a more specific population with CKD in more advanced stages (3b-4).

### Patient and Public Involvement

There was no direct patient and public involvement in the design, conduct, or reporting of this study.

### Sample size

We did not conduct a formal sample size calculation, as we had access to all data collected routinely. Nevertheless, we were cognisant of the unreliability of performance assessment with insufficient sample sizes, particularly in the context of a low number of events. Consequently, we restricted our analysis to clinically relevant subpopulations, ensuring that each group contained a minimum of 100 events and non-events [27]. Given these constraints, we deemed it unreliable to analyse groups 3a, 3b, and 4 separately. Instead, we combined them into subgroups 3a-3b-4 and 3b-4 for the analysis.

### Validation Model

Tangri et al. developed the KFRE model in Canada in 2011 to predict kidney failure in populations with CKD stages 3 to 5 [23]. The model was later recalibrated based on an extensive meta-analysis that included 31 cohorts from more than 30 countries and over 72,000 participants [30]. The 4-variable KFRE includes age, patient sex, eGFR, and ACR collected on the same date for each patient and is an attractive alternative since it requires few variables that are easily accessible in the Peruvian health system. This version has two prediction horizons: short-term at 2 years and long-term at 5 years (equations in **Table S1**).

### Predictors

The four predictors of the 4-variable KFRE are age (years), sex (male/female), eGFR (ml/min/1.73m^2^), and ACR (mg/g) (see **Table S2**). eGFR was calculated using the 2009 CKD Epidemiology Collaboration (CKD-EPI) formula [20,31], considering patient ethnicity, which clinicians ascertained through direct observation and recorded solely as black/non-black for eGFR calculation purposes (see **Supplementary Methods – section 1.3** for details). The health establishments in the Rebagliati Network have standardized care protocols for CKD patients, including laboratory procedures, as part of the Peruvian National Kidney Health Plan. Serum creatinine levels for eGFR estimation were determined from a blood sample. For ACR calculation, urine creatinine and albumin levels were determined by quantitative and automated laboratory tests from a random urine sample. Qualified personnel verified the preanalytical conditions in each hospital. The urine samples were collected in 10-15 ml bottles and transported between 4 and 8 °C to the respective laboratory for daily processing. The entire analytical process followed good laboratory and analytical quality control practices. We obtained all data on these variables from UMERC, a computer app designed specifically for this purpose.

### Outcome variable

The outcome variable was kidney failure, defined as KRT-treated end-stage renal disease (ESRD), which in Peru comprises the initiation of hemodialysis or peritoneal dialysis indicated by a nephrologist and based on clinical parameters of uremia and eGFR < 15ml/min/1.73m^2^.

To estimate the observed risk of kidney failure, we considered the competing event of death without KRT. The data on the date of admission to KRT were obtained from the dialysis database and were verified in the digital clinical history. The date of death until December 31, 2019, was obtained from the National Registry of Identification and Civil Status (RENIEC) of Peru.

### Follow-up time

We followed patients until they developed kidney failure, death, or censorship, whichever occurred first. We censored observations until the patient was lost to follow-up or the end of the study (December 31, 2019). We chose to end the study on this date to avoid having data from the pandemic, during which the health system collapsed, kidney care services were affected, and the reliability of the information was altered.

### Statistical analysis

#### Initial data analysis

We performed an initial data analysis to identify implausible extreme values, missing data, and inconsistencies. Plausible extreme data was retained without any transformation in the main analysis. Numerical and categorical variables were described using median (interquartile ranges) and absolute frequencies (percentages), respectively.

#### Estimate of observed risk

We estimated non-parametric cumulative incidence function (CFI) curves and their 95% confidence intervals (95% CI) using the Aalen-Johansen estimator [32] for kidney failure and considering the competing risk of death without kidney failure.

#### Predictive Performance of KFRE

We estimated the individual predicted risks of developing kidney failure using the 4-variable KFRE for 2- and 5-year horizons (prediction formulas in **Table S1**). We assessed the performance of the models based on discrimination and calibration measures [27,28] according to TRIPOD guidelines. Additionally, we considered the risk of death without kidney failure and based our analysis workflow on two recently published methodological guides on external validation of prediction models in the presence of competing risks [33,34].

Discrimination is a relative measure of how well the model distinguishes between patients with or without the condition of interest [34,35]. To assess discrimination, we estimated the truncated concordance index (C-index) at 2- and 5-years for each model and the areas under the ROC time-dependent curves of cumulative sensitivity and dynamic specificity (C/D AUC-td) [36]. A C-index or C/D AUC-td of 1 indicates perfect discrimination, 0.5 indicates no discrimination, and values ≥ 0.8 are generally considered appropriate for prognostic models [35]. We accounted for the competing risk for death without kidney failure by censoring patients who die at infinite, indicating that they may not develop kidney failure in the future [33,34].

Calibration is a measure that indicates how well the absolute predicted risks agree with the observed risks. These observed risks were estimated using CFI to takte into account the competing risk of death without kidney failure [33,34]. We assessed calibration-in-the-large using the observed to expected results (O/E), a measure of mean calibration, and the calibration intercept, a measure of weak calibration. We also assessed weak calibration through calibration slope. Moderate calibration was assessed inspecting calibration plots.

We estimated the ratio of observed to expected results (O/E). An O/E indicates perfect global calibration, an O/E > 1 indicates an underestimation of the average risk, and an O/E < 1 reveals an overestimated of the average risk. The calibration intercept is another measure that evaluates the average over or underestimation that we estimate in this study. An intercept of 0 indicates perfect agreement between the predicted and observed risk average. An intercept significantly < 0 indicates an overestimation, and an intercept > 0 indicates an underestimation of the risk average. We also estimated the calibration slope. A slope of 1 reflects ideal agreement. A slope less than 1 indicates that the predicted risks are too extreme (very high and low), while a slope greater than 1 indicates that the predictions do not show enough variation. To formally test statistical evidence of miscalibration, we first performed a Wald test of the joint contribution of the intercept and slope, as previously described for calibration models in prediction models [33,34].

Calibration plots allow calibration to be assessed in detail by comparing observed individual risks with those predicted. A curve exactly following the 45° straight line would indicate a perfect situation named strong calibration that is ideal and utopic. A more realistic goal is to assess if the curve is close to the diagonal indicating moderate calibration. We plotted calibration curves estimated by smoothed local linear regression (loess) based on pseudovalues obtained from cumulative incidence estimates that account for the competing risk of death [33,34,37].

#### Sensitivity analysis

We perform two sensitivity analyses:

1. The same analysis approach after eliminating the extreme values of ACR by winzoring at the 1st and 99th percentiles of the distribution of this variable.
2. A predictive performance analysis ignoring competitive risk. This analysis relied extensively on the methodology described by McLernon DJ et al [37].

#### General approach

The data preparation and all the analyzes were carried out with the statistical program R version 4.2.1 for Windows 11 x 64 bits. As there were no missing data in critical variables, all analysis were case-complete. Except for the C-index, all the 95% confidence interval (CI) were Wald-type [33,34]. The 95% CI for C-index was obtained using the percentile bootstrap method using 1000 bootstrapped samples [33,34].

## RESULTS

### Study population

Of the 22,745 patients screened from 2013 to 2017 at 17 Lima hospitals, 7,519 with CKD stages 3a-4 and 2,798 with CKD stages 3b-4 were eligible, having complete ACR data (**Figure 1**). All eligible patients had complete data on age, sex, and outcome. The median observation time was 4.9 years in the CKD 3a-4 group.

**Figure 1.**
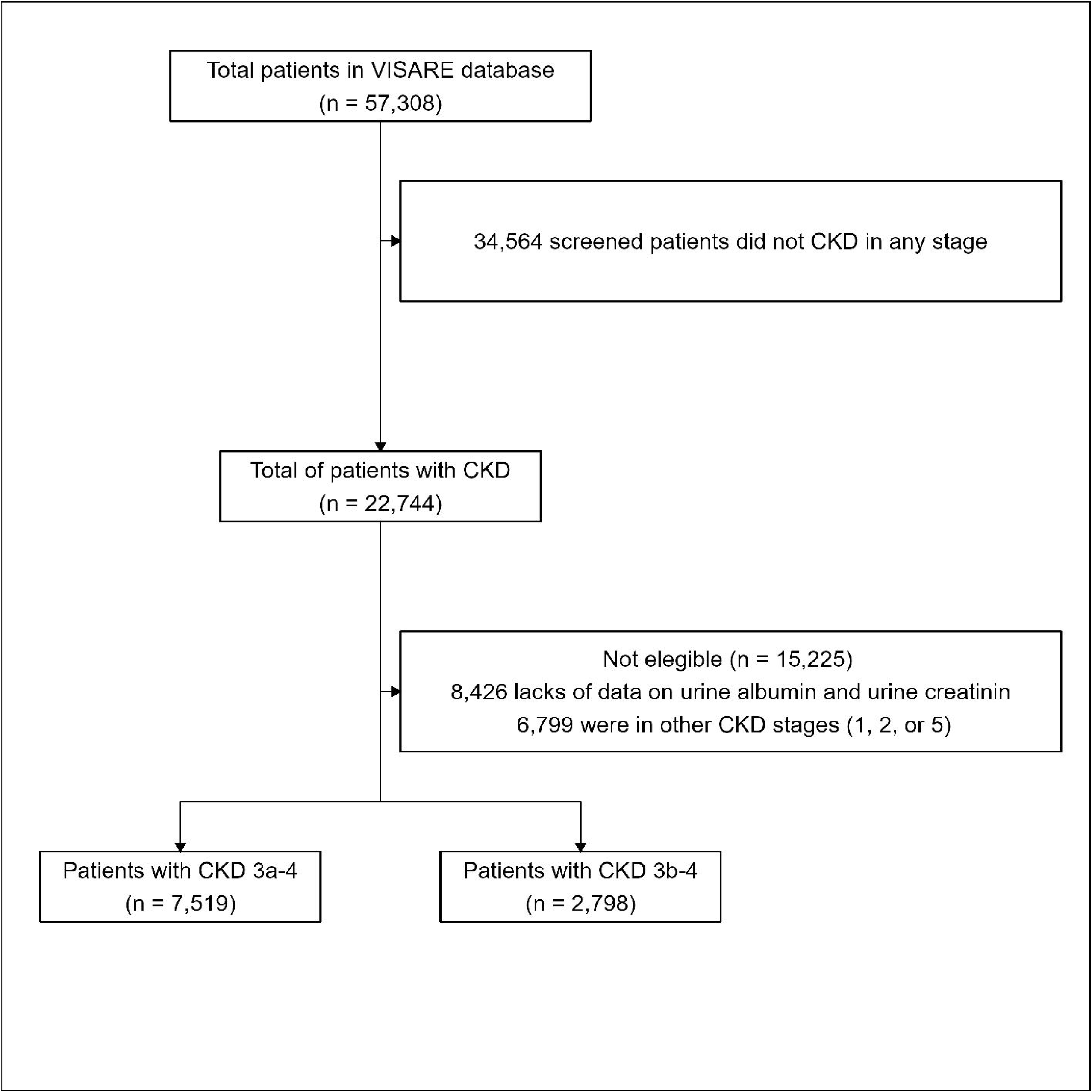
Study flowchart.

**Table 1** outlines the baseline characteristics of the study population, while Supplementary Results present the characteristics of the study population according to kidney failure at 2 and 5 years for CKD 3a-4 (**Table S3**) and CKD 3b-4 (**Table S4**) subgroups, respectively. Kidney failure case numbers at 2 years were low for subpopulations with stages 3a (n=26), 3b (n=36), and 4 (n=52) (**Table S5**), and similarly low for 5-year case numbers in stages 3a (n=57), 3b (n=81), and 4 (n=101) (**Table S5**). Consequently, evaluating predictive performance in these specific subgroups was deemed unreliable. The distribution of patients in stages 3a-4 and 3b-4 across the EsSalud Rebagliati Network’s 17 health facilities is detailed in supplementary material **Tables S6** and **S7**.

**Table 1.** Baseline characteristics of the study population according to CKD stages.

| **Characteristic** | **CKD Stages 3a-3b-4**<br>**(n = 7,519)** | **CKD Stages 3b-4**<br>**(n = 2,798)** |
| --- | --- | --- |
| **Sex** |  |  |
| Female | 4,107 (54.6%) | 1,398 (50.0%) |
| Male | 3,412 (45.4%) | 1,400 (50.0%) |
| **Age (years)** |  |  |
| Mean (SD) | 74.0 (10.2) | 75.6 (10.6) |
| Median (IQR) | 75.0 (68.0 - 81.0) | 77.0 (70.0 - 83.0) |
| Range | 23.0 - 97.0 | 23.0 - 97.0 |
| **Ethnicity** |  |  |
| Black | 0 (0%) | 0 (0%) |
| Non-black | 7,519 (100%) | 2,798 (100%) |
| **Hypertension** | 4,486 (59.7%) | 1,636 (58.5%) |
| **Diabetes Mellitus** | 1,845 (24.5%) | 674 (24.1%) |
| **Persistent albuminuria categories** |  |  |
| A1 | 4,772 (63.5%) | 1,494 (53.4%) |
| A2 | 2,018 (26.8%) | 860 (30.7%) |
| A3 | 729 (9.7%) | 444 (15.9%) |
| **GFR categories** |  |  |
| G3a | 4,721 (62.8%) |  |
| G3b | 2,207 (29.4%) | 2,207 (78.9%) |
| G4 | 591 (7.9%) | 591 (21.1%) |
| **CKD KDIGO classification** |  |  |
| Moderately increased risk | 3,278 (43.6%) |  |
| High risk | 2,460 (32.7%) | 1,302 (46.5%) |
| Very high risk | 1,781 (23.7%) | 1,496 (53.5%) |
| **Serum Creatinine (mg/dL)** |  |  |
| Mean (SD) | 1.4 (0.4) | 1.7 (0.5) |
| Median (IQR) | 1.3 (1.1 - 1.5) | 1.6 (1.4 - 1.9) |
| Range | 0.8 - 3.9 | 1.1 - 3.9 |
| **eGFR (ml/min/1.73m<sup>2</sup>)** |  |  |
| Mean (SD) | 46.2 (9.8) | 35.7 (7.3) |
| Median (IQR) | 48.7 (40.4 - 53.8) | 37.3 (31.4 - 41.7) |
| Range | 15.0 - 60.0 | 15.0 - 45.0 |
| **ACR (mg/g)** |  |  |
| Mean (SD) | 248.6 (3,044.4) | 334.1 (3,050.8) |
| Median (IQR) | 14.6 (4.5 - 66.1) | 26.0 (6.5 - 153.8) |
| Range | 0.0 - 144,870.6 | 0.0 - 144,870.6 |
| **Urine Albumin (mg/ml)** |  |  |
| Mean (SD) | 8.3 (28.3) | 13.1 (36.3) |
| Median (IQR) | 1.0 (0.3 - 4.0) | 1.7 (0.4 - 10.1) |
| Range | 0.0 - 658.0 | 0.0 - 658.0 |
| **Urine Creatinine (mg/dl)** |  |  |
| Mean (SD) | 72.4 (47.5) | 71.4 (47.0) |
| Median (IQR) | 63.3 (41.3 - 86.0) | 64.9 (43.3 - 85.0) |
| Range | 0.1 - 722.1 | 0.7 - 722.1 |
| **Death at 2 years\*** | 640 (8.5%) | 358 (12.8%) |
| **Outcome at 2 years** |  |  |
| Alive w/o Kidney Failure | 6,842 (91.0%) | 2,410 (86.1%) |
| Death w/o Kidney Failure | 563 (7.5%) | 300 (10.7%) |
| Kidney Failure | 114 (1.5%) | 88 (3.1%) |
| **Death at 5 years\*** | **1,539 (20.5%)** | **784 (28.0%)** |
| **Outcome at 5 years** |  |  |
| Alive w/o Kidney Failure | 5,880 (78.2%) | 1,933 (69.1%) |
| Death w/o Kidney Failure | 1,400 (18.6%) | 683 (24.4%) |
| Kidney Failure | 239 (3.2%) | 182 (6.5%) |

SD: standard deviation, IQR: interquartile range, ACR: urine albumin to creatinine ratio, eGFR: glomerular filtration rate estimated by CKD-EPI formula
\* Death after or before kidney failure

### Observed and predicted risk of kidney failure

**Figure 2** displays the observed risk of kidney failure and death without kidney failure for both study populations. The 2- and 5-year observed risks of kidney failure in patients with CKD stages 3a-4 were 1.52% and 3.37%, respectively (**Table S7**). In patients with CKD 3b-4, the 2- and 5-year observed risks of kidney failure were 3.15% and 6.87%, respectively (**Table S8**). The distribution of the 2- and 5-year predicted risk by KFRE are shown in **Figure S1**.

**Figure 2.**
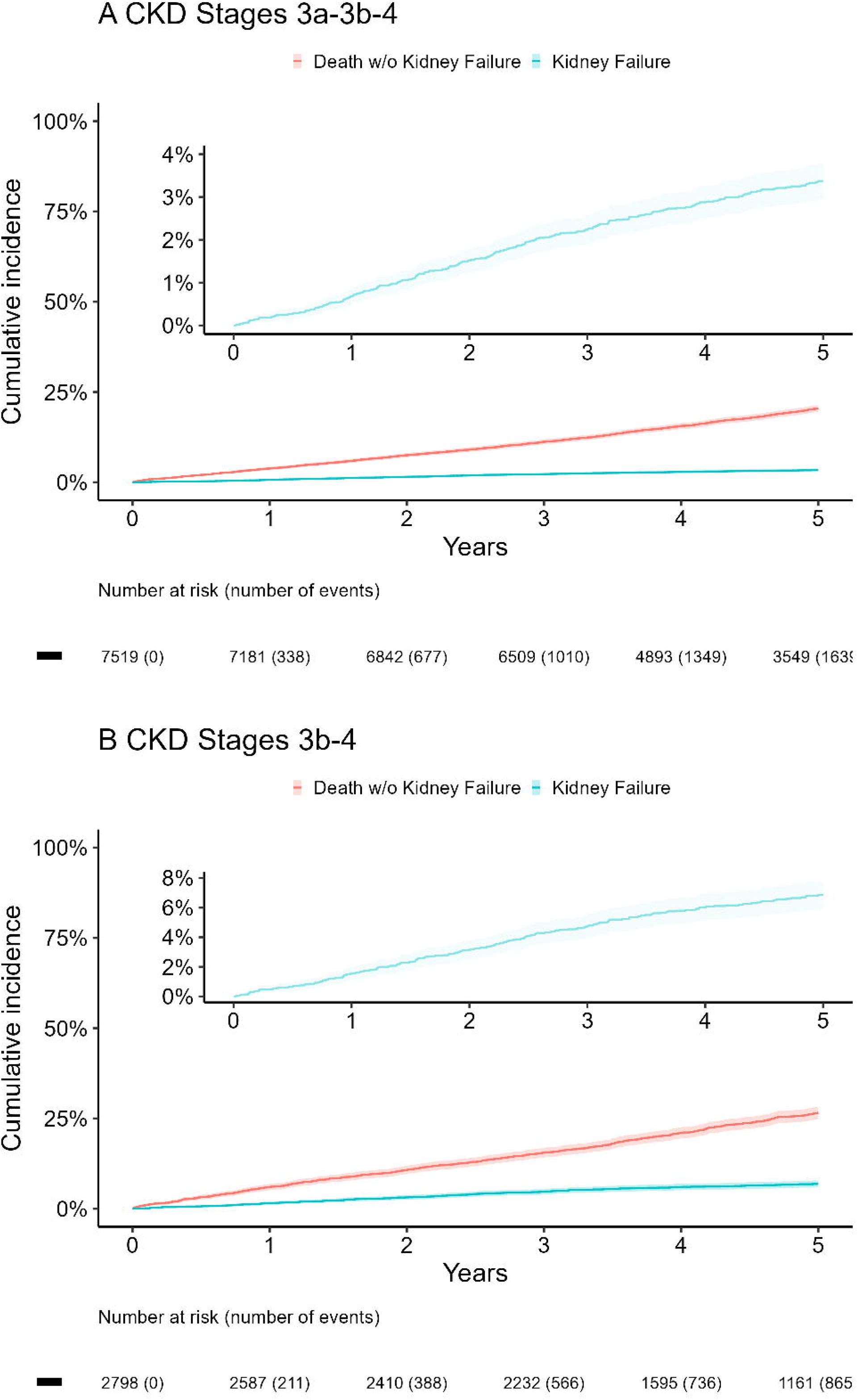
Cumulative incidence function curves for kidney failure (sky-blue line) and death before kidney failure (red line) in patients with (A) CKD stages 3a-3b-4 and (B) CKD stages 3b-4. CKD: Chronic Kidney Disease.

### KFRE predictive performance

KFRE demonstrated good discriminatory ability across all time horizons and study populations, as evidenced by C-index and C/D AUC-td values exceeding 0.8 (**Table 2**). However, miscalibration tests indicate low compatibility with good calibration of KFRE at all-time horizons and groups (all p-values ≤0.001) (**Table 2**).

**Table 2.** Performance measures of KFRE in the external dataset of patients with CKD stages 3a-3b-4 and 3b-4.

| Validation aspect and performance measure | CKD Stages 3a-3b-4 |  | CKD Stages 3b-4 |  |
| --- | --- | --- | --- | --- |
|  | t = 2 year | t = 5 year | t = 2 year | t = 5 year |
| **Calibration** |  |  |  |  |
| Average predicted risk | 0.96% | 3.18% | 2.36% | 7.66% |
| Average observed risk (95% CI) | 1.52% (1.24% to 1.79%) | 3.37% (2.95% to 3.8%) | 3.15% (2.5% to 3.79%) | 6.86% (5.89% to 7.84%) |
| O/E ratio (95% CI) | 1.57 (1.39 to 1.76) | 1.06 (0.93 to 1.19) | 1.33 (1.13 to 1.54) | 0.9 (0.75 to 1.04) |
| Calibration intercept (95% CI) | 0.18 (-0.1 to 0.45) | -0.26 (-0.45 to -0.07) | 0.16 (-0.12 to 0.44) | -0.29 (-0.48 to -0.1) |
| Calibration slope (95% CI) | 0.79 (0.61 to 0.96) | 0.75 (0.65 to 0.86) | 0.82 (0.6 to 1.03) | 0.79 (0.66 to 0.92) |
| **Discrimination** |  |  |  |  |
| C-index up to t-years (95% CI) | 0.853 (0.812 to 0.892) | 0.845 (0.818 to 0.872) | 0.848 (0.803 to 0.885) | 0.827 (0.796 to 0.857) |
| C/D AUC, at t years (95% CI) | 0.855 (0.816 to 0.895) | 0.847 (0.819 to 0.875) | 0.853 (0.811 to 0.895) | 0.836 (0.803 to 0.869) |

CKD: chronic kidney disease, t: time, O/E: Observed versus expected outcome ratio %: percentage, 95%CI: 95% confidence interval, C-index: truncated agreement index, C/D AUC-td: area under ROC curves time dependent on cumulative sensitivity and dynamic specificity.

Regarding calibration in the large, for patients with CKD stages 3a-4, the 2-year average observed risk of kidney failure was 1.52%, while KFRE predicted a lower average risk at 0.96%, yielding an O/E ratio of 1.57, indicating an overall 2-year risk underestimation. A similar pattern was observed for CKD stages 3b-4 patients (O/E ratio: 1.33). In contrast, the imprecision of calibration intercepts hindered the evaluation of KFRE’s 2-year calibration in-the-large.

For the 5-year KFRE model, evidence of poor calibration in the large was also observed, although in the opposite direction, suggesting an overprediction. The O/E ratio was less useful for evaluating long-term KFRE calibration due to high imprecision, but calibration intercepts showed overestimation of 5-year risk for both CKD stages 3a-3b-4 (calibration intercept: -0.26) and CKD stages 3b-4 subgroups (calibration intercept: -0.29) (**Table 2**).

Poor weak calibration was also evident at 2-years, with extreme predictions for CKD stages 3a-3b-4 (calibration slope: 0.79) and CKD stages 3b-4 (calibration slope: 0.82). The 5-year KFRE also showed extreme predictions for both groups, with calibration slopes of 0.75 for CKD stages 3a-3b-4 and 0.79 for CKD stages 3b-4 (**Table 2**).

Regarding evidence of poor moderate calibration, calibration curves (Figure 3) displayed 2-year risk underestimation mainly in patients with predicted risk less than 0.3-0.4 for both CKD stages 3a-4 and CKD stages 3b-4 subgroups (Figures 3A and 3C). In contrast, 5-year risk overestimation mainly occurred in individuals with predicted risk greater than 0.2 for both CKD stages 3a-4 subgroups.

**Figure 3.**
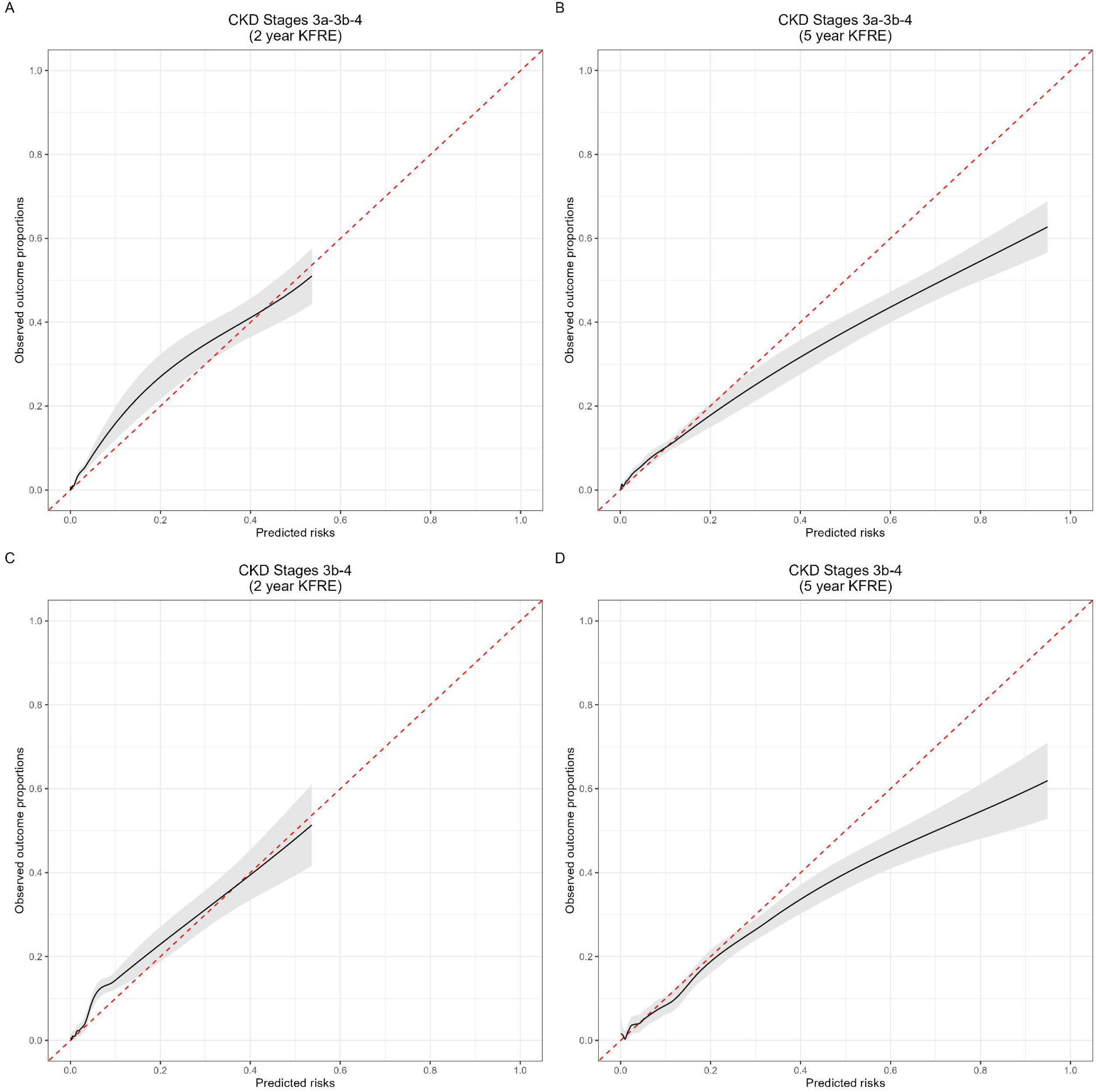
Calibration curves for each group and prediction horizon. The x-axis shows the risk predicted by the KFRE model, and the y-axis represents the observed risk estimated using the cumulative incidence function to consider the competing risk of death without kidney failure. CKD: Chronic Kidney Disease.

### Sensitivity analysis: Impact of outliers in ACR

The distribution of the four KFRE equation variables and the distribution of risks predicted by KFRE are shown in **Figure S1** and **Figure S2**, respectively. We noted very extreme values in the ACR variable (**Figure S2C** and **Figure S2D**), leading to a sensitivity analysis assessing the robustness of our predictive performance after mitigating these extreme values.

Winsorisation of ACR’s extreme values was applied at its 1st and 99th percentiles, and risks predicted by KFRE were recalculated using the winsorised ACR variable. Despite notable changes in ACR’s distribution after winsorisation (**Figure S3**), the distribution of risks predicted by KFRE in the original data remained largely unaltered (**Figure S4**). Median and mean values of risks predicted by KFRE before and after winsorisation were strikingly similar, as were variability measures such as standard deviation, interquartile range, and range (**Table S10**). The predictive performance of KFRE on the winsorised data closely resembled the original data (**Table S11** and **Figure S5**).

### Sensitivity analysis: Predictive performance assessment ignoring competing risk

We evaluated the predictive performance results when not accounting for competing risks. We discovered that the 2-year incidence of kidney failure in CKD stages 3a-4, without considering competing risk, was 1.58%, only marginally higher than when accounting for competing risk (1.52%) (**Fig S6**). At 5 years, these differences became more pronounced but remained relatively small, with a 3.37% incidence when considering competing risk and 4.24% when not considering competing risk. In the CKD stages 3b-4 population, the 5-year differences were approximately 2% (6.89% when considering competing risk vs. 8.99% when not considering competing risk) (**Fig S6**). Thus, the assessment of KFRE’s predictive performance without considering competing risks also identified KFRE as miscalibrated (**Table S12** and **Fig S6**); however, the magnitude of miscalibration was substantially smaller when ignoring competing risks compared to when considering competing risks (**Fig S7**).

## DISCUSSION

### Principal findings

We independently validated the 4-variable KFRE for kidney failure prognosis at 2- and 5-years in Peruvian patients with CKD stages 3a-4 and 3b-4. Although KFRE demonstrated good discrimination, its calibration was poor. The model underestimated average short-term risk (2-years) and overestimated average long-term risk (5-years) of kidney failure in CKD stages 3a-4 patients. This pattern persisted in the CKD stages 3b-4 subgroup. KFRE also had poor weak calibration manifested as very extreme predictions, while poor moderate calibration was evident in the underestimation of actual short-term risk in patients with KFRE-predicted risks below 0.3-0.4 and overestimation of individual long-term risks, primarily in individuals with a KFRE-predicted risk greater than 0.2.

### Comparison with previous literature

The KFRE has been externally validated in numerous global studies, mainly in North American [23,30,39,41,43–48], European [16,30,40,49], and Asian countries [38,42]. However, few validations have been conducted in Latin America, particularly at the primary care level [30]. Existing validations in Latin American may not accurately represent current CKD populations due to advancements in CKD management [30]. This highlights the need for updated KFRE validation studies in Latin America.

In line with prior research, the non-North American versions of the KFRE models at 2- and 5-year intervals exhibited good discrimination in our study and across diverse population groups from countries [16,30,38,40,42,49]. The initial external validation of the non-North American KFRE, a meta-analysis comprising 13 cohorts including Chilean and Brazilian patients [30], reported pooled C-statistics of 0.9 and 0.88 for predicting 2- and 5-year kidney failure, respectively [30]. A more recent study observed lower discrimination values, with C-indexes ranging from 0.76-0.84 for 2-year KFRE and 0.75-0.81 for 5-year KFRE in cohorts from Germany, Italy, the Netherlands, Poland, Switzerland, and the United Kingdom, none of which included Latin American countries [16]. Despite the lower values in the recent study, the discrimination remained good. These findings are consistent with other studies reporting C-index values greater than 0.8 for the non-North American KFRE at 2- and 5-years [38,40,42,49].

In contrast to discrimination, our study’s calibration assessment results differ from the initial study that recalibrated and validated KFRE for non-North American populations [30]. Although the few studies that assess calibration of the non-North American version of KFRE have reported poor calibration [33,40,42,49], most primarily focus on moderate calibration using calibration curves, with limited attention given to calibration in the large or weak calibration. Our findings for moderate calibration are in line with previous studies that identified overprediction of kidney failure risk at 5 years [33,40,42,49], particularly in individuals with high predicted risk groups (>0.3 to 0.4) [42,49].

On the other hand, the 2-year results display greater heterogeneity among existing studies: some cohorts show good calibration [33,42], others overpredict risk in high- and low-risk groups [40]. In our study, we found that KFRE exhibits an opposite pattern of underprediction at 2 years. Regarding calibration in the large, only Ramspek et al. assessed this aspect, finding an overprediction of the average 5-year risk by >10% while observing good calibration in the large at 2 years. This contrasts with our study, which also identified underestimation of the actual average risk by KFRE at 2 years in patients with CKD 3a-4 and CKD 3b-4.

Differences in CKD severity or stages between the initial study validation and our investigation may account for these discrepancies. In our study, for instance, over half of the patients were classified as having moderate CKD severity (stage G3a), and we excluded patients with advanced CKD (stage G5) (**Table 1**). In contrast, Tangri et al. incorporated stage 5 CKD patients; however, it is challenging to evaluate the impact of these differences as stage distribution information was not reported by Tangri et al. We also found no significant differences in eGFR and albuminuria distributions, which are recognised prognostic markers for kidney failure in CKD patients (**Table S13**). The initial study reported mean (standard deviation) eGFR values of 47 ml/min/1.73m2 (12 ml/min/1.73m2) and a 34% prevalence of albuminuria for their non-North American population, while our study reported similar values of 46.2 ml/min/1.73m2 (9.8 ml/min/1.73m2) and 36.5%, respectively (**Table S13**). The actual event risk may also explain the model’s miscalibration. Kidney failure incidence rates suggest a lower risk of kidney failure in our study’s CKD stages 3a-4 patients (7.4 per 1000 person-years) compared to 9.2 per 1000 person-years.

Another explanation for the pattern of overprediction we found in the long term for the KFRE model, especially in advanced CKD populations [43–45], is partly due to not accounting for death without kidney failure as a competing risk [33,34]. This competing risk is crucial for patients with advanced CKD, especially in frail or older populations requiring long-term predictions with more frequent death events [50]. Most existing models censor patients who die, leading to overestimation of the actual risk [51]. Ramspek et al. [16] and Ravani et al. [52] found that KFRE overestimated the actual average risk of terminal CKD by 10-18% and 1-27% at 5 years, respectively, with overestimation increasing over time among high-risk individuals atributable to competing event. By this reason, we considered the competing risk of death without kidney failure in our study. Initial study validating KFRE for non-North American populations also evaluated the impact of competing risk but found no significant differences.

In our study involving patients with CKD stages 3a-4, 7.5% died without kidney failure at 2 years of follow-up, and 20.5% at 5 years (**Table S8**). Conversely, the cumulative incidence of kidney failure was low, with 1.5% at two years and 3.4% at 5 years. This demonstrates the relatively minor impact of competing risk at 2 years, which becomes substantially more significant at 5 years. Even without considering competing risk, the Cox analysis revealed miscalibration, displaying the same patterns as the competing risk analysis, although the degree of miscalibration would have been less pronounced.

It is important to note that, while differences exist between using Cox and competing risk analyses at the 2-year horizon, these disparities are minimal in our study. In contrast, at 5 years, marked differences emerge, with the Cox analysis evidently biasing the performance evaluation. Therefore, we chose to report the competing risk analysis as our primary method and the Cox analysis as secondary. This approach better reflects the increasing impact of competing risk when the incidence of the competing event (death without renal failure) becomes more frequent, as observed in the 5-year assessment.

### Strengths and limitations of this study

The present study offers the first external validation of KFRE in Peru, utilising a retrospective cohort of over 7,000 patients from 17 primary and secondary care EsSalud health establishments in Lima. Furthermore, as far as we know, this investigation serves as the second study of the Kidney Failure Risk Equation (KFRE) in Latin America, boasting several strengths. The prior research was a meta-analysis validating the original equation for non-North American populations, including cohorts from Chile and Brazil [30].

The study employs robust statistical methods and sound analytical techniques for assessing KFRE performance, considering the competing risk of death without renal failure. This approach helps avoid overestimation of the observed risk and reduces bias in performance assessment [33,34,50,53]. The decision to use this competing risk approach as a primary analysis, with Cox methods employed in a sensitivity analysis, was informed by contemporary evidence demonstrating that accounting for competing risks offers a less biased and more accurate estimation of the actual kidney failure risks [33,52]. As such, this study adheres to best practices in the field and contributes essential knowledge regarding KFRE’s performance in Peru and Latin America more broadly.

Although our study presents valuable findings; it is important to acknowledge several limitations. Firstly, we utilized secondary data routinely recorded by multiple evaluating clinicians across 17 healthcare centers in Lima. On one hand, using routine clinical data has the advantage of potentially reflecting the model’s performance more accurately in real-world clinical practice. However, despite standardization of laboratory measurements as part of the National Kidney Program in Peru, clinical registries are inherently susceptible to errors in data recording, thereby introducing the potential for measurement error.

Another limitation stems from our use of RRT initiation as an indicator of kidney failure in patients. This approach carries the risk of misclassification for patients who may have opted for conservative management. Nevertheless, it is worth noting that conservative management is relatively rare in Peru. Furthermore, access to RRT for EsSalud beneficiaries in Lima is fully covered by EsSalud, which bears the entire cost reducing the risk of not access to RRT, specially in Lima. These factors imply that the risk of bias might be reduced; nevertheless, we still regard this as a significant limitation that should be appropriately addressed in future prospective studies.

Lastly, although the similarities among EsSalud service networks in Lima may support the notion that our findings could be generalizable to other networks in the city, it is crucial to recognize that Lima is not representative of the entirety of Peru, and EsSalud is not the sole healthcare system in the country [54,55]. Significant disparities exist in healthcare services provided outside Lima and between different healthcare systems, such as Comprehensive Health Insurance and the Health of the Armed and Police Forces. These variations may influence KFRE’s predictive performance, necessitating specific external validation studies in these populations due to the distinct differences among Peruvian health subsystems.

### Implications for clinical practice

The observed differences in KFRE model performance among various cohorts highlight the necessity of broadening external validation across diverse populations and settings [25]. Although our study demonstrates KFRE’s capacity to effectively discriminate between patients who will develop kidney failure at 2 and 5 years, it also reveals the model’s shortcomings in accurately predicting individual risks, underestimating them in the short term and overestimating them in the long term. Sole reliance on KFRE’s discrimination ability may have adverse implications for patients. Given our findings that short-term overestimation of kidney failure risk occurs in patients with low predicted risk and long-term underestimation occurs in those with high predicted risk, this pattern of poor moderate calibration could result in detrimental patient outcomes.

For instance, if KFRE overestimates the risk of kidney failure in patients with a lower true progression risk, this could result in unnecessary referrals for dialysis preparation, which may cause undue distress to patients and potentially increase the risk of death from preventable cardiovascular events if patients had remained under primary care. Conversely, if KFRE underestimates the risk of kidney failure in patients with a higher true progression risk, it could lead to delays in their referral and preparation for dialysis.

### Future research

Future research should focus on model updating for clinically relevant populations in Peru in a national scale. It is also essential to examine the influence of patient mix heterogeneity on model performance and propose specific updates for enhanced local performance [33]. Moreover, the clinical utility of KFRE and its impact on significant patient and healthcare system outcomes should be evaluated.

### Conclusions

The KFRE model exhibited good discrimination at 2 and 5 years for patients with CKD stages 3a-3b-4 and CKD stages 3b-4. However, the model was found to be miscalibrated, underestimating short-term risks and overestimating long-term risks. Despite its considerable discriminative capacity, the KFRE model requires updating before being recommended for use in clinical practice guidelines for the Peruvian population. In light of this evidence, Latin American countries should consider externally validating and updating this model prior to recommending its use in clinical practice.

## Supporting information

Supplemental Material TRIPOD Checklist

Supplemental Material Methods and Results

## Data Availability

The analysis code essential for the replication of study findings can be accessed at this link: https://github.com/psotob91/kfre-ckd-reba-peru. As per the privacy policies of EsSalud, the minimal data set is not open to public access. However, we are amenable to providing the anonymised data upon receipt of a reasonable request directed to the corresponding author.

## ACKNOWLEDGMENTS

The authors would like to acknowledge the methodological advisoring of the Instituto de Evaluación de Tecnologías en Salud e Investigación – IETSI, during the initial stages of the project, through its Personalised Mentoring Programme 2020, in the development of this study.

## AUTHOR CONTRIBUTORS

JBZ served as the principal investigator, conceived the study concept, developed the proposal, contributed to the study design, coordinated the project, and participated in drafting the manuscript. RCG co-authored the proposal, contributed to the study design, oversaw data acquisition and preprocessing, and assisted in drafting the manuscript. PSB co-authored the proposal, contributed to the study design, was responsible for cleaning the raw data, conducting the analysis, and preparing the manuscript. JBZ and PSB serve as guarantors, ensuring the integrity of the study. The corresponding author attests that all listed authors meet authorship criteria and that no others meeting the criteria have been omitted.

## ETHICAL APPROVAL

The Research Ethics Committee of the Edgardo Rebagliati National Hospital approved this study (575-GRPR-ESSALUD-2021). Because all the data from clinical registries were anonymized before being processed for the research, the ethics committee accepted the waiver of informed consent from the patients.

## TRANSPARENCY DECLARATION

JBZ and PSB, as the manuscript’s guarantors, affirm that the manuscript presents an honest, accurate, and transparent account of the study reported. They confirm that no significant aspects of the study have been omitted, and any discrepancies from the planned study have been duly explained.

