## Supplemental Material TRIPOD Checklist for "Kidney Failure Prediction: Multicenter External Validation of KFRE Model in Patients with CKD Stages 3-4 in Peru"

TRIPOD Checklist: Prediction Model Development and Validation

| Section/Topic | Item | | Checklist Item | Page |
|---|---|---|---|---|
| **Title and abstract** | | | | |
| Title | 1 | D;V | Identify the study as developing and/or validating a multivariable prediction model, the target population, and the outcome to be predicted. | 1 |
| Abstract | 2 | D;V | Provide a summary of objectives, study design, setting, participants, sample size, predictors, outcome, statistical analysis, results, and conclusions. | 2 |
| **Introduction** | | | | |
| Background and objectives | 3a | D;V | Explain the medical context (including whether diagnostic or prognostic) and rationale for developing or validating the multivariable prediction model, including references to existing models. | 4-5 |
| | 3b | D;V | Specify the objectives, including whether the study describes the development or validation of the model or both. | 5 |
| **Methods** | | | | |
| Source of data | 4a | D;V | Describe the study design or source of data (e.g., randomized trial, cohort, or registry data), separately for the development and validation data sets, if applicable. | 5 |
| | 4b | D;V | Specify the key study dates, including start of accrual; end of accrual; and, if applicable, end of follow-up. | 5, 7 |
| Participants | 5a | D;V | Specify key elements of the study setting (e.g., primary care, secondary care, general population) including number and location of centres. | 5 |
| | 5b | D;V | Describe eligibility criteria for participants. | 5 |
| | 5c | D;V | Give details of treatments received, if relevant. | NA |
| Outcome | 6a | D;V | Clearly define the outcome that is predicted by the prediction model, including how and when assessed. | 7 |
| | 6b | D;V | Report any actions to blind assessment of the outcome to be predicted. | NA |
| Predictors | 7a | D;V | Clearly define all predictors used in developing the multivariable prediction model, including how and when they were measured. | 6 |
| | 7b | D;V | Report any actions to blind assessment of predictors for the outcome and other predictors. | NA |
| Sample size | 8 | D;V | Explain how the study size was arrived at. | 6 |
| Missing data | 9 | D;V | Describe how missing data were handled (e.g., complete-case analysis, single imputation, multiple imputation) with details of any imputation method. | 9 |
| Statistical analysis methods | 10a | D | Describe how predictors were handled in the analyses. | NA |
| | 10b | D | Specify type of model, all model-building procedures (including any predictor selection), and method for internal validation. | NA |
| | 10c | V | For validation, describe how the predictions were calculated. | 6-7 |
| | 10d | D;V | Specify all measures used to assess model performance and, if relevant, to compare multiple models. | 8-9 |
| | 10e | V | Describe any model updating (e.g., recalibration) arising from the validation, if done. | NA |
| Risk groups | 11 | D;V | Provide details on how risk groups were created, if done. | NA |
| Development vs. validation | 12 | V | For validation, identify any differences from the development data in setting, eligibility criteria, outcome, and predictors. | Supplementary Material, Table S13 (page 17) |
| **Results** | | | | |
| Participants | 13a | D;V | Describe the flow of participants through the study, including the number of participants with and without the outcome and, if applicable, a summary of the follow-up time. A diagram may be helpful. | 9 |
| | 13b | D;V | Describe the characteristics of the participants (basic demographics, clinical features, available predictors), including the number of participants with missing data for predictors and outcome. | 9-10 |
| | 13c | V | For validation, show a comparison with the development data of the distribution of important variables (demographics, predictors and outcome). | Supplementary Material, Table S13 (page 17) |
| Model development | 14a | D | Specify the number of participants and outcome events in each analysis. | NA |
| | 14b | D | If done, report the unadjusted association between each candidate predictor and outcome. | NA |
| Model specification | 15a | D | Present the full prediction model to allow predictions for individuals (i.e., all regression coefficients, and model intercept or baseline survival at a given time point). | NA |
| | 15b | D | Explain how to use the prediction model. | NA |
| Model performance | 16 | D;V | Report performance measures (with CIs) for the prediction model. | 10-12 |
| Model-updating | 17 | V | If done, report the results from any model updating (i.e., model specification, model performance). | NA |
| **Discussion** | | | | |
| Limitations | 18 | D;V | Discuss any limitations of the study (such as nonrepresentative sample, few events per predictor, missing data). | 16-17 |
| Interpretation | 19a | V | For validation, discuss the results with reference to performance in the development data, and any other validation data. | 13-15 |
| | 19b | D;V | Give an overall interpretation of the results, considering objectives, limitations, results from similar studies, and other relevant evidence. | 13-18 |
| Implications | 20 | D;V | Discuss the potential clinical use of the model and implications for future research. | 17-18 |
| **Other information** | | | | |
| Supplementary information | 21 | D;V | Provide information about the availability of supplementary resources, such as study protocol, Web calculator, and data sets. | Supplementary PDF and Code in Github repo |
| Funding | 22 | D;V | Give the source of funding and the role of the funders for the present study. | 19 |

### TRIPOD Checklist: Prediction Model Development and Validation

*Items relevant only to the development of a prediction model are denoted by D, items relating solely to a validation of a prediction model are denoted by V, and items relating to both are denoted D;V. We recommend using the TRIPOD Checklist in conjunction with the TRIPOD Explanation and Elaboration document.
