## Supplemental Material Methods and Results for "Kidney Failure Prediction: Multicenter External Validation of KFRE Model in Patients with CKD Stages 3-4 in Peru"

**SUPPLEMENTARY MATERIAL**

### 1. SUPPLEMENTARY METHODS

#### 1.1. KFRE equations validated externally by this study

This study performed external validation of the recalibrated equations for the non-North American population, which are available on page nine of the study supplementary material by *Tangri N, et al* (1) and were recommended as preferable by the article. These equations are implemented in the following web app: https://kidneyfailurerisk.com/. The equations we validated are shown in **Table S1**.

**Table S1. KFRE equations externally validated by the study**

| Prediction horizons | Original regional equation calibrated for predicted risk of kidney failure |
|---|---|
| 2-years | $1 - 0.9832^{e^{(-0.2201\times(\frac{age}{10}-7.036)+0.2467\times(male-0.5642)-0.5567\times(\frac{eGFR}{5}-7.222)+0.4510\times(\log(ACR)-5.13}}}$ |
| 5-years | $1 - 0.9365^{e^{(-0.2201\times(\frac{age}{10}-7.036)+0.2467\times(male-0.5642)-0.5567\times(\frac{eGFR}{5}-7.222)+0.4510\times(\log(ACR)-5.13}}}$ |

*Source: Tangri N, et al. (1)*

#### 1.2. Variables

**Table S2. Coding of variables**

| Variable | Coding |
|---|---|
| age | integer number that indicates the age in completed years |
| male | 1 = male; 0 = female |
| eGFR_ckdepi | estimated glomerular filtration rate obtained by CKD-EPI formula in ml/min/1.73m$^2$ |
| acr | albumin-to-creatinine ratio in mg/g |

#### 1.3. 2009 CKD Epidemiology Collaboration (CKD-EPI) formula for eGFR

$$eGFR = A \times (SCr/B)^{C} \times 0.993^{age} \times (1.159\ if\ black)$$

Where $A, B, and\ C$ are the following:

| Female | | Male | |
|---|---|---|---|
| $SCr \leq 0.7$ | $A = 144$<br>$B = 0.7$<br>$C = -0.329$ | $SCr \leq 0.9$ | $A = 141$<br>$B = 0.9$<br>$C = -0.411$ |
| $SCr > 0.7$ | $A = 144$<br>$B = 0.7$<br>$C = -1.209$ | $SCr > 0.9$ | $A = 141$<br>$B = 0.9$<br>$C = -1.209$ |

#### 2. SUPPLEMENTARY TABLES

**Table S3. Baseline characteristics of study patients at CKD Stages 3a, 3b, or 4 for all predictors, stratified by 2- and 5-year outcomes.**

| | 2-years | | 5-years | |
|---|---|---|---|---|
| **Characteristic** | **No kidney failure (n = 7,405)** | **Kidney failure (n = 114)** | **No kidney failure (n = 7,280)** | **Kidney failure (n = 239)** |
| **Sex** | | | | |
| Female | 4,062 (54.9%) | 45 (39.5%) | 4,015 (55.2%) | 92 (38.5%) |
| Male | 3,343 (45.1%) | 69 (60.5%) | 3,265 (44.8%) | 147 (61.5%) |
| **Age (years)** | | | | |
| Mean (SD) | 74.1 (10.2) | 66.6 (11.7) | 74.2 (10.1) | 66.5 (12.5) |
| Median (IQR) | 75.0 (68.0, 81.0) | 67.0 (59.2, 74.0) | 75.0 (68.0, 82.0) | 67.0 (59.0, 75.0) |
| Range | 23.0, 97.0 | 36.0, 88.0 | 23.0, 97.0 | 26.0, 94.0 |
| **Hypertension** | 4,421 (59.7%) | 65 (57.0%) | 4,339 (59.6%) | 147 (61.5%) |
| **Diabetes Mellitus** | 1,796 (24.3%) | 49 (43.0%) | 1,743 (23.9%) | 102 (42.7%) |
| **Persistent albuminuria categories** | | | | |
| A1 | 4,749 (64.1%) | 23 (20.2%) | 4,725 (64.9%) | 47 (19.7%) |
| A2 | 1,984 (26.8%) | 34 (29.8%) | 1,941 (26.7%) | 77 (32.2%) |
| A3 | 672 (9.1%) | 57 (50.0%) | 614 (8.4%) | 115 (48.1%) |
| **GFR categories** | | | | |
| G3a | 4,695 (63.4%) | 26 (22.8%) | 4,664 (64.1%) | 57 (23.8%) |
| G3b | 2,171 (29.3%) | 36 (31.6%) | 2,126 (29.2%) | 81 (33.9%) |
| G4 | 539 (7.3%) | 52 (45.6%) | 490 (6.7%) | 101 (42.3%) |
| **CKD KDIGO classification** | | | | |
| Moderately increased risk | 3,266 (44.1%) | 12 (10.5%) | 3,256 (44.7%) | 22 (9.2%) |
| High risk | 2,448 (33.1%) | 12 (10.5%) | 2,429 (33.4%) | 31 (13.0%) |
| Very high risk | 1,691 (22.8%) | 90 (78.9%) | 1,595 (21.9%) | 186 (77.8%) |
| **Serum Creatinine (mg/dL)** | | | | |
| Mean (SD) | 1.4 (0.4) | 2.0 (0.7) | 1.3 (0.4) | 2.0 (0.7) |
| Median (IQR) | 1.3 (1.1, 1.5) | 1.9 (1.5, 2.5) | 1.3 (1.1, 1.5) | 1.9 (1.5, 2.4) |
| Range | 0.8, 3.8 | 1.0, 3.9 | 0.8, 3.8 | 0.9, 3.9 |
| **eGFR (ml/min/1.73m2)** | | | | |
| Mean (SD) | 46.4 (9.6) | 33.7 (12.5) | 46.6 (9.4) | 34.5 (12.4) |

| | | | | |
|---|---|---|---|---|
| Median (IQR) | 48.9 (40.8, 53.9) | 31.7 (23.2, 42.8) | 49.0 (41.1, 53.9) | 33.0 (23.5, 44.2) |
| Range | 15.0, 60.0 | 15.4, 59.8 | 15.1, 60.0 | 15.0, 59.8 |
| **ACR (mg/g)** | | | | |
| Mean (SD) | 235.5 (3,059.4) | 1,101.7 (1,614.1) | 229.3 (3,083.4) | 836.1 (1,283.1) |
| Median (IQR) | 14.2 (4.4, 62.7) | 302.0 (52.5, 1,663.9) | 14.0 (4.4, 59.4) | 270.9 (51.2, 992.4) |
| Range | 0.0, 144,870.6 | 2.5, 7,462.7 | 0.0, 144,870.6 | 0.2, 7,462.7 |
| **Urine Albumin (mg/ml)** | | | | |
| Mean (SD) | 7.6 (26.5) | 52.1 (73.5) | 7.1 (25.7) | 44.0 (62.3) |
| Median (IQR) | 0.9 (0.3, 3.7) | 16.0 (4.3, 71.6) | 0.9 (0.3, 3.5) | 15.3 (3.4, 56.5) |
| Range | 0.0, 658.0 | 0.2, 348.1 | 0.0, 658.0 | 0.0, 348.1 |
| **Urine Creatinine (mg/dl)** | | | | |
| Mean (SD) | 72.5 (47.7) | 64.8 (33.7) | 72.4 (47.0) | 69.8 (61.0) |
| Median (IQR) | 63.4 (41.4, 86.5) | 59.6 (39.6, 85.0) | 63.5 (41.3, 86.6) | 58.7 (41.0, 85.0) |
| Range | 0.1, 722.1 | 6.4, 218.6 | 0.1, 620.1 | 6.4, 722.1 |
| **Death at 2 years*** | 563 (7.6%) | 77 (67.5%) | 563 (7.7%) | 77 (32.2%) |
| **Outcome at 2 years** | | | | |
| Alive w/o Kidney Failure | 6,842 (92.4%) | 0 (0.0%) | 6,717 (92.3%) | 125 (52.3%) |
| Death w/o Kidney Failure | 563 (7.6%) | 0 (0.0%) | 563 (7.7%) | 0 (0.0%) |
| Kidney Failure | 0 (0.0%) | 114 (100.0%) | 0 (0.0%) | 114 (47.7%) |
| **Death at 5 years*** | 1,462 (19.7%) | 77 (67.5%) | 1,400 (19.2%) | 139 (58.2%) |
| **Outcome at 5 years** | | | | |
| Alive w/o Kidney Failure | 5,880 (79.4%) | 0 (0.0%) | 5,880 (80.8%) | 0 (0.0%) |
| Death w/o Kidney Failure | 1,400 (18.9%) | 0 (0.0%) | 1,400 (19.2%) | 0 (0.0%) |
| Kidney Failure | 125 (1.7%) | 114 (100.0%) | 0 (0.0%) | 239 (100.0%) |

IQR: interquartile range, ACR: urine albumin to creatinine ratio, eGFR: glomerular filtration rate estimated by CKD-EPI formula

* Death after or before kidney failure

**Table S4. Baseline characteristics of study patients at CKD Stages 3b or 4 for all predictors, stratified by 2- and 5-year outcomes.**

| | 2-years | | 5-years | |
|---|---|---|---|---|
| **Characteristic** | **No kidney failure (n = 2,710)** | **Kidney failure (n = 88)** | **No kidney failure (n = 2,616)** | **Kidney failure (n = 182)** |
| **Sex** | | | | |
| Female | 1,363 (50.3%) | 35 (39.8%) | 1,329 (50.8%) | 69 (37.9%) |
| Male | 1,347 (49.7%) | 53 (60.2%) | 1,287 (49.2%) | 113 (62.1%) |
| **Age (years)** | | | | |
| Mean (SD) | 75.9 (10.4) | 66.1 (12.1) | 76.2 (10.2) | 66.9 (12.4) |
| Median (IQR) | 77.0 (70.0, 83.0) | 67.0 (59.0, 74.0) | 77.0 (70.0, 83.0) | 67.0 (59.2, 75.0) |
| Range | 23.0, 97.0 | 36.0, 88.0 | 23.0, 97.0 | 26.0, 94.0 |
| **Hypertension** | 1,581 (58.3%) | 55 (62.5%) | 1,517 (58.0%) | 119 (65.4%) |
| **Diabetes Mellitus** | 635 (23.4%) | 39 (44.3%) | 596 (22.8%) | 78 (42.9%) |
| **Persistent albuminuria categories** | | | | |
| A1 | 1,483 (54.7%) | 11 (12.5%) | 1,469 (56.2%) | 25 (13.7%) |
| A2 | 830 (30.6%) | 30 (34.1%) | 798 (30.5%) | 62 (34.1%) |
| A3 | 397 (14.6%) | 47 (53.4%) | 349 (13.3%) | 95 (52.2%) |
| **GFR categories** | | | | |
| G3a | 0 (0.0%) | 0 (0.0%) | 0 (0.0%) | 0 (0.0%) |
| G3b | 2,171 (80.1%) | 36 (40.9%) | 2,126 (81.3%) | 81 (44.5%) |
| G4 | 539 (19.9%) | 52 (59.1%) | 490 (18.7%) | 101 (55.5%) |
| **CKD KDIGO classification** | | | | |
| Moderately increased risk | 0 (0.0%) | 0 (0.0%) | 0 (0.0%) | 0 (0.0%) |
| High risk | 1,294 (47.7%) | 8 (9.1%) | 1,286 (49.2%) | 16 (8.8%) |
| Very high risk | 1,416 (52.3%) | 80 (90.9%) | 1,330 (50.8%) | 166 (91.2%) |
| **Serum Creatinine (mg/dL)** | | | | |
| Mean (SD) | 1.7 (0.4) | 2.3 (0.6) | 1.7 (0.4) | 2.2 (0.6) |
| Median (IQR) | 1.6 (1.4, 1.9) | 2.1 (1.8, 2.5) | 1.6 (1.4, 1.8) | 2.1 (1.7, 2.5) |
| Range | 1.1, 3.8 | 1.2, 3.9 | 1.1, 3.8 | 1.2, 3.9 |
| **eGFR (ml/min/1.73m$^2$)** | | | | |
| Mean (SD) | 35.9 (7.1) | 28.2 (8.1) | 36.1 (7.0) | 29.0 (8.3) |
| Median (IQR) | 37.6 (31.8, 41.8) | 26.9 (21.6, 34.9) | 37.8 (32.1, 41.9) | 28.3 (22.4, 35.4) |
| Range | 15.0, 45.0 | 15.4, 43.8 | 15.1, 45.0 | 15.0, 44.9 |
| **ACR (mg/g)** | | | | |
| Mean (SD) | 306.8 (3,082.4) | 1,172.8 (1,631.1) | 296.0 (3,133.9) | 881.4 (1,272.2) |
| Median (IQR) | 24.6 (6.3, 145.4) | 367.7 (149.1, 1,811.8) | 23.3 (6.1, 131.2) | 334.6 (143.8, 1,076.5) |

| | | | | |
|---|---|---|---|---|
| Range | 0.0, 144,870.6 | 2.5, 7,462.7 | 0.0, 144,870.6 | 1.4, 7,462.7 |
| **Urine Albumin (mg/ml)** | | | | |
| Mean (SD) | 11.7 (33.3) | 56.1 (77.1) | 10.7 (32.2) | 47.4 (64.4) |
| Median (IQR) | 1.5 (0.4, 8.7) | 16.3 (8.9, 66.6) | 1.4 (0.4, 7.7) | 16.5 (8.6, 60.3) |
| Range | 0.0, 658.0 | 0.2, 348.1 | 0.0, 658.0 | 0.1, 348.1 |
| **Urine Creatinine (mg/dl)** | | | | |
| Mean (SD) | 71.7 (47.4) | 62.1 (33.5) | 71.7 (45.8) | 68.3 (61.8) |
| Median (IQR) | 65.1 (43.7, 85.0) | 55.3 (38.1, 85.0) | 65.4 (43.9, 85.0) | 58.0 (40.1, 85.0) |
| Range | 0.7, 722.1 | 6.4, 218.6 | 0.7, 620.1 | 6.4, 722.1 |
| **Death at 2 years*** | 300 (11.1%) | 58 (65.9%) | 300 (11.5%) | 58 (31.9%) |
| **Outcome at 2 years** | | | | |
| Alive w/o Kidney Failure | 2,410 (88.9%) | NA | 2,316 (88.5%) | 94 (51.6%) |
| Death w/o Kidney Failure | 300 (11.1%) | NA | 300 (11.5%) | NA |
| Kidney Failure | NA | 88 (100.0%) | NA | 88 (48.4%) |
| **Death at 5 years*** | 726 (26.8%) | 58 (65.9%) | 683 (26.1%) | 101 (55.5%) |
| **Outcome at 5 years** | | | | |
| Alive w/o Kidney Failure | 1,933 (71.3%) | 0 (0.0%) | 1,933 (73.9%) | 0 (0.0%) |
| Death w/o Kidney Failure | 683 (25.2%) | 0 (0.0%) | 683 (26.1%) | NA |
| Kidney Failure | 94 (3.5%) | 88 (100.0%) | NA | 182 (100.0%) |

IQR: interquartile range, ACR: urine albumin to creatinine ratio, eGFR: glomerular filtration rate estimated by CKD-EPI formula. NA: not applicable

* Death after or before kidney failure

**Table S5. Baseline characteristics and frequency of outcomes in the study population according to CKD stages.**

| Characteristic | G3a (n = 4,721) | G3b (n = 2,207) | G4 (n = 591) |
|---|---|---|---|
| **Sex** | | | |
| Female | 2,709 (57.4%) | 1,102 (49.9%) | 296 (50.1%) |
| Male | 2,012 (42.6%) | 1,105 (50.1%) | 295 (49.9%) |
| **Age (years)** | | | |
| Mean (SD) | 73.1 (9.9) | 76.0 (10.2) | 73.9 (11.8) |
| Median (IQR) | 74.0 (67.0 - 80.0) | 77.0 (70.0 - 83.0) | 75.0 (67.0 - 83.0) |
| Range | 23.0 - 97.0 | 26.0 - 97.0 | 23.0 - 95.0 |
| **Hypertension** | 2,850 (60.4%) | 1,295 (58.7%) | 341 (57.7%) |
| **Diabetes Mellitus** | 1,171 (24.8%) | 521 (23.6%) | 153 (25.9%) |
| **Persistent albuminuria categories** | | | |
| A1 | 3,278 (69.4%) | 1,302 (59.0%) | 192 (32.5%) |
| A2 | 1,158 (24.5%) | 632 (28.6%) | 228 (38.6%) |
| A3 | 285 (6.0%) | 273 (12.4%) | 171 (28.9%) |
| **CKD KDIGO classification** | | | |
| Moderately increased risk | 3,278 (69.4%) | 0 (0.0%) | 0 (0.0%) |
| High risk | 1,158 (24.5%) | 1,302 (59.0%) | 0 (0.0%) |
| Very high risk | 285 (6.0%) | 905 (41.0%) | 591 (100.0%) |
| **Serum Creatinine (mg/dL)** | | | |
| Mean (SD) | 1.2 (0.2) | 1.5 (0.2) | 2.3 (0.5) |
| Median (IQR) | 1.1 (1.0 - 1.3) | 1.5 (1.3 - 1.7) | 2.3 (1.9 - 2.6) |
| Range | 0.8 - 1.9 | 1.1 - 2.4 | 1.5 - 3.9 |
| **eGFR (ml/min/1.73m$^2$)** | | | |
| Mean (SD) | 52.5 (3.9) | 38.8 (4.2) | 24.2 (4.1) |
| Median (IQR) | 52.6 (49.4 - 55.6) | 39.3 (35.3 - 42.4) | 24.7 (21.2 - 27.7) |
| Range | 45.0 - 60.0 | 30.0 - 45.0 | 15.0 - 30.0 |
| **ACR (mg/g)** | | | |
| Mean (SD) | 198.0 (3,039.8) | 302.6 (3,386.3) | 451.7 (1,109.7) |
| Median (IQR) | 10.8 (3.9 - 41.5) | 20.3 (5.3 - 100.1) | 127.2 (17.7 - 405.2) |
| Range | 0.0 - 137,672.1 | 0.0 - 144,870.6 | 0.1 - 18,259.2 |
| **Urine Albumin (mg/ml)** | | | |
| Mean (SD) | 5.5 (21.8) | 10.0 (31.4) | 25.0 (48.8) |
| Median (IQR) | 0.7 (0.2 - 2.4) | 1.3 (0.3 - 5.8) | 7.4 (1.3 - 20.9) |
| Range | 0.0 - 548.0 | 0.0 - 658.0 | 0.0 - 383.0 |
| **Urine Creatinine (mg/dl)** | | | |
| Mean (SD) | 72.9 (47.9) | 71.3 (46.0) | 72.0 (50.7) |
| Median (IQR) | 62.4 (40.4 - 88.1) | 64.8 (43.0 - 85.0) | 65.3 (44.6 - 85.0) |
| Range | 0.1 - 438.3 | 0.8 - 620.1 | 0.7 - 722.1 |
| **Death at 2 years*** | 282 (6.0%) | 230 (10.4%) | 128 (21.7%) |
| **Outcome at 2 years** | | | |
| Alive w/o Kidney Failure | 4,432 (93.9%) | 1,965 (89.0%) | 445 (75.3%) |
| Death w/o Kidney Failure | 263 (5.6%) | 206 (9.3%) | 94 (15.9%) |
| Kidney Failure | 26 (0.6%) | 36 (1.6%) | 52 (8.8%) |

| | | | |
|---|---|---|---|
| **Death at 5 years*** | 755 (16.0%) | 555 (25.1%) | 229 (38.7%) |
| **Outcome at 5 years** | | | |
| Alive w/o Kidney Failure | 3,947 (83.6%) | 1,622 (73.5%) | 311 (52.6%) |
| Death w/o Kidney Failure | 717 (15.2%) | 504 (22.8%) | 179 (30.3%) |
| Kidney Failure | 57 (1.2%) | 81 (3.7%) | 101 (17.1%) |

IQR: interquartile range, ACR: urine albumin to creatinine ratio, eGFR: glomerular filtration rate estimated by CKD-EPI formula

* Death after or before kidney failure

**Table S6. Distribution of CKD 3a-4 patients included in the analysis across 17 health facilities of the EsSalud Rebagliati Network**

| | | Outcome | | |
|---|---|---|---|---|
| **Healthcare center*** | **Overall (n = 7,519)** | **Alive w/o Kidney Failure (n = 5,880)** | **Death w/o Kidney Failure (n = 1,400)** | **Kidney Failure (n = 239)** |
| Healthcare №1 | 74 (100.0%) | 48 (64.9%) | 18 (24.3%) | 8 (10.8%) |
| Healthcare №2 | 58 (100.0%) | 52 (89.7%) | 6 (10.3%) | 0 (0.0%) |
| Healthcare №3 | 403 (100.0%) | 319 (79.2%) | 73 (18.1%) | 11 (2.7%) |
| Healthcare №4 | 287 (100.0%) | 204 (71.1%) | 71 (24.7%) | 12 (4.2%) |
| Healthcare №5 | 308 (100.0%) | 242 (78.6%) | 53 (17.2%) | 13 (4.2%) |
| Healthcare №6 | 1,471 (100.0%) | 1,088 (74.0%) | 298 (20.3%) | 85 (5.8%) |
| Healthcare №7 | 34 (100.0%) | 20 (58.8%) | 11 (32.4%) | 3 (8.8%) |
| Healthcare №8 | 1,032 (100.0%) | 858 (83.1%) | 160 (15.5%) | 14 (1.4%) |
| Healthcare №9 | 207 (100.0%) | 158 (76.3%) | 42 (20.3%) | 7 (3.4%) |
| Healthcare №10 | 258 (100.0%) | 179 (69.4%) | 73 (28.3%) | 6 (2.3%) |
| Healthcare №11 | 77 (100.0%) | 71 (92.2%) | 5 (6.5%) | 1 (1.3%) |
| Healthcare №12 | 1,289 (100.0%) | 1,058 (82.1%) | 202 (15.7%) | 29 (2.2%) |
| Healthcare №13 | 679 (100.0%) | 497 (73.2%) | 166 (24.4%) | 16 (2.4%) |
| Healthcare №14 | 752 (100.0%) | 586 (77.9%) | 143 (19.0%) | 23 (3.1%) |
| Healthcare №15 | 454 (100.0%) | 381 (83.9%) | 64 (14.1%) | 9 (2.0%) |
| Healthcare №16 | 102 (100.0%) | 90 (88.2%) | 10 (9.8%) | 2 (2.0%) |
| Healthcare №17 | 34 (100.0%) | 29 (85.3%) | 5 (14.7%) | 0 (0.0%) |

* For privacy reasons, the name of the current healthcare provider is not disclosed. However, we can provide the names internally if the request is reasonable and appropriate mechanisms are in place to ensure the protection of data privacy.

**Table S7. Distribution of CKD 3b-4 patients included in the analysis across 17 health facilities of the EsSalud Rebagliati Network.**

| | | Outcome | | |
|---|---|---|---|---|
| **Healthcare center*** | **Overall (n = 2,798)** | **Alive w/o Kidney Failure (n = 1,933)** | **Death w/o Kidney Failure (n = 683)** | **Kidney Failure (n = 182)** |
| Healthcare №1 | 26 (100.0%) | 10 (38.5%) | 10 (38.5%) | 6 (23.1%) |
| Healthcare №2 | 12 (100.0%) | 8 (66.7%) | 4 (33.3%) | 0 (0.0%) |
| Healthcare №3 | 129 (100.0%) | 87 (67.4%) | 32 (24.8%) | 10 (7.8%) |
| Healthcare №4 | 113 (100.0%) | 68 (60.2%) | 35 (31.0%) | 10 (8.8%) |
| Healthcare №5 | 115 (100.0%) | 81 (70.4%) | 24 (20.9%) | 10 (8.7%) |
| Healthcare №6 | 921 (100.0%) | 644 (69.9%) | 212 (23.0%) | 65 (7.1%) |
| Healthcare №7 | 22 (100.0%) | 11 (50.0%) | 8 (36.4%) | 3 (13.6%) |
| Healthcare №8 | 271 (100.0%) | 194 (71.6%) | 67 (24.7%) | 10 (3.7%) |
| Healthcare №9 | 70 (100.0%) | 47 (67.1%) | 17 (24.3%) | 6 (8.6%) |
| Healthcare №10 | 90 (100.0%) | 59 (65.6%) | 28 (31.1%) | 3 (3.3%) |
| Healthcare №11 | 15 (100.0%) | 13 (86.7%) | 1 (6.7%) | 1 (6.7%) |
| Healthcare №12 | 408 (100.0%) | 294 (72.1%) | 93 (22.8%) | 21 (5.1%) |
| Healthcare №13 | 216 (100.0%) | 134 (62.0%) | 70 (32.4%) | 12 (5.6%) |
| Healthcare №14 | 215 (100.0%) | 152 (70.7%) | 47 (21.9%) | 16 (7.4%) |
| Healthcare №15 | 144 (100.0%) | 106 (73.6%) | 30 (20.8%) | 8 (5.6%) |
| Healthcare №16 | 23 (100.0%) | 18 (78.3%) | 4 (17.4%) | 1 (4.3%) |
| Healthcare №17 | 8 (100.0%) | 7 (87.5%) | 1 (12.5%) | 0 (0.0%) |

* For privacy reasons, the name of the current healthcare provider is not disclosed. However, we can provide the names internally if the request is reasonable and appropriate mechanisms are in place to ensure the protection of data privacy.

**Table S8. Cumulative incidence of kidney failure and death without kidney failure in patients with CKD stages 3a-3b-4.**

| Year | Kidney failure | | Death without kidney failure | |
|---|---|---|---|---|
| | % | 95% CI | % | 95% CI |
| 1-year | 0.68% | (0.49% to 0.86%) | 3.82% | (3.38% to 4.25%) |
| 2-year | 1.52% | (1.24% to 1.79%) | 7.49% | (6.89% to 8.08%) |
| 3-year | 2.23% | (1.9% to 2.57%) | 11.2% | (10.48% to 11.91%) |
| 4-year | 2.88% | (2.5% to 3.26%) | 15.58% | (14.75% to 16.41%) |
| 5-year | 3.37% | (2.95% to 3.8%) | 20.46% | (19.48% to 21.42%) |

%: observed risk estimated utilising the cumulative incidence function with the Aalen-Johansen estimator to account for competing risk; CI: confidence interval

**Table S9. Cumulative incidence of kidney failure and death without kidney failure in patients with CKD stages 3b-4.**

| Year | Kidney failure | | Death without kidney failure | |
|---|---|---|---|---|
| | % | 95% CI | % | 95% CI |
| 1-year | 1.54% | (1.08% to 1.99%) | 6% | (5.12% to 6.88%) |
| 2-year | 3.15% | (2.5% to 3.79%) | 10.72% | (9.57% to 11.86%) |
| 3-year | 4.72% | (3.93% to 5.5%) | 15.51% | (14.16% to 16.84%) |
| 4-year | 6.02% | (5.12% to 6.9%) | 21.03% | (19.48% to 22.55%) |
| 5-year | 6.86% | (5.89% to 7.83%) | 26.59% | (24.84% to 28.3%) |

%: observed risk estimated utilising the cumulative incidence function with the Aalen-Johansen estimator to account for competing risk; CI: confidence interval; CI: confidence interval

**Table S10. Summary of ACR, 2-year and 5-year predicted risks of kidney failure according to KFRE before and after winsorising the 1% and 99% extreme values of ACR.**

| | **Original data** | **Winsorised outliers in ACR.** |
|---|---|---|
| **Characteristic** | **N = 7,519** | **N = 7,519** |
| **ACR (mg/g)** | | |
| Mean ± SD | 248.620 ± 3,044.427 | 144.053 ± 437.405 |
| Median (IQR) | 14.634 (4.484, 66.114) | 14.634 (4.484, 66.114) |
| Range | 0.002, 144,870.588 | 0.236, 3,195.727 |
| **Predicted risk of kidney failure to 2 years (%)** | | |
| Mean ± SD | 0.963 ± 3.263 | 0.944 ± 3.155 |
| Median (IQR) | 0.131 (0.057, 0.429) | 0.131 (0.057, 0.429) |
| Range | 0.001, 53.657 | 0.006, 53.657 |
| **Predicted risk of kidney failure to 5 years (%)** | | |
| Mean ± SD | 3.182 ± 8.913 | 3.138 ± 8.760 |
| Median (IQR) | 0.507 (0.220, 1.650) | 0.508 (0.221, 1.649) |
| Range | 0.004, 94.911 | 0.024, 94.911 |

SD: standard deviation, IQR: interquartile range, ACR: urine albumin to creatinine ratio

**Table S11. Performance measures of KFRE in the external dataset of patients with CKD stages 3a-3b-4 and 3b-4 following the application of winsorisation to the ACR variable and the subsequent recalculation of predicted risks using the Kidney Failure Risk Equation (KFRE).**

| Validation aspect and performance measure | CKD Stages 3a-3b-4 | | CKD Stages 3b-4 | |
|---|---|---|---|---|
| | t = 2 year | t = 5 year | t = 2 year | t = 5 year |
| **Calibration** | | | | |
| Average predicted risk | 0.94% | 3.14% | 2.32% | 7.58% |
| Average observed proportion (95% CI) | 1.52% (1.24% to 1.79%) | 3.37% (2.95% to 3.8%) | 3.15% (2.5% to 3.79%) | 6.86% (5.89% to 7.84%) |
| O/E ratio (95% CI) | 1.61 (1.42 to 1.79) | 1.08 (0.95 to 1.2) | 1.35 (1.15 to 1.56) | 0.91 (0.76 to 1.05) |
| Calibration intercept (95% CI) | 0.21 (-0.06 to 0.49) | -0.23 (-0.41 to -0.04) | 0.19 (-0.09 to 0.47) | -0.26 (-0.45 to -0.07) |
| Calibration slope (95% CI) | 0.79 (0.61 to 0.97) | 0.77 (0.66 to 0.87) | 0.82 (0.61 to 1.04) | 0.8 (0.67 to 0.94) |
| **Discrimination** | | | | |
| C-index up to t-years (95% CI) | 0.853 (0.812 to 0.893) | 0.845 (0.818 to 0.872) | 0.848 (0.804 to 0.885) | 0.828 (0.797 to 0.857) |
| C/D AUC, at t years (95% CI) | 0.855 (0.816 to 0.895) | 0.848 (0.82 to 0.875) | 0.854 (0.812 to 0.896) | 0.837 (0.804 to 0.87) |

CKD: chronic kidney disease, t: time, O/E: Observed versus expected outcome ratio %: percentage, 95%CI: 95% confidence interval, C-index: truncated agreement index, C/D AUC-td: area under ROC curves time dependent on cumulative sensitivity and dynamic specificity.

**Table S12. Performance measures of KFRE in the external dataset of patients with CKD stages 3a-3b-4 and 3b-4, without considering competing risks.**

| Validation aspect and performance measure | CKD Stages 3a-3b-4 | | CKD Stages 3b-4 | |
|---|---|---|---|---|
| | t = 2 year | t = 5 year | t = 2 year | t = 5 year |
| **Calibration** | | | | |
| Average predicted risk | 0.96% | 3.18% | 2.36% | 7.66% |
| Average observed proportion | 1.58% | 3.72% | 3.35% | 7.88% |
| O/E ratio (95% CI) | 1.64 (1.37 to 1.97) | 1.17 (1.03 to 1.33) | 1.42 (1.15 to 1.75) | 1.03 (0.89 to 1.19) |
| Calibration slope (95% CI) | 0.79 (0.72 to 0.85) | 0.79 (0.72 to 0.85) | 0.83 (0.74 to 0.93) | 0.83 (0.74 to 0.93) |
| **Discrimination** | | | | |
| Harrell C (95% CI) | 0.856 (0.817 to 0.895) | 0.856 (0.83 to 0.883) | 0.85 (0.809 to 0.891) | 0.839 (0.809 to 0.869) |
| Uno C (95% CI) | 0.856 (0.817 to 0.895) | 0.851 (0.825 to 0.878) | 0.851 (0.81 to 0.892) | 0.832 (0.801 to 0.864) |
| C/D AUC, at t years (95% CI) | 0.861 (0.822 to 0.9) | 0.86 (0.832 to 0.887) | 0.86 (0.818 to 0.901) | 0.846 (0.812 to 0.879) |

CKD: chronic kidney disease, t: time, O/E: Observed versus expected outcome ratio %: percentage, 95%CI: 95% confidence interval, C/D AUC-td: area under ROC curves time dependent on cumulative sensitivity and dynamic specificity.

**Table S13. Comparisons of characteristics of original cohort that recalibrated Non North American version of KFRE and our study population**

| | | Current study | |
|---|---|---|---|
| **Characteristics** | **Original Study (Non-North American population)** | **CKD Stages 3a-4** | **CKD Stages 3b-4** |
| Numbef of participants | 103753 | 7519 | 2798 |
| F/U Time, years, Median (IQR) | 4 (3, 6) | 4.9 (3.5, 5.9) | 4.6 (3.2, 5.8) |
| Age, years (SD) | 71 (12) | 74 (10.2) | 75.6 (10.6) |
| Male, n (%) | 46632 (45%) | 3412 (45.4%) | 1400 (50%) |
| Black ethnicity, n (%) | 393 (0.4%) | 0 (0%) | 0 (0%) |
| eGFR, ml/min/1.73m$^2$ (SD) | 47 (12) | 46.2 (9.8) | 35.7 (7.3) |
| Albuminuria, n (%) | 24962 (34%) | 2747 (36.5%) | 1304 (46.6%) |
| Kidney Failure Incidence (per 1000 py) | 9.2 | 7.4 | 16.1 |

F/U: follow-up; eGFR: glomerular filtration rate estimated by CKD-EPI formula

#### 3. SUPPLEMENTARY FIGURES

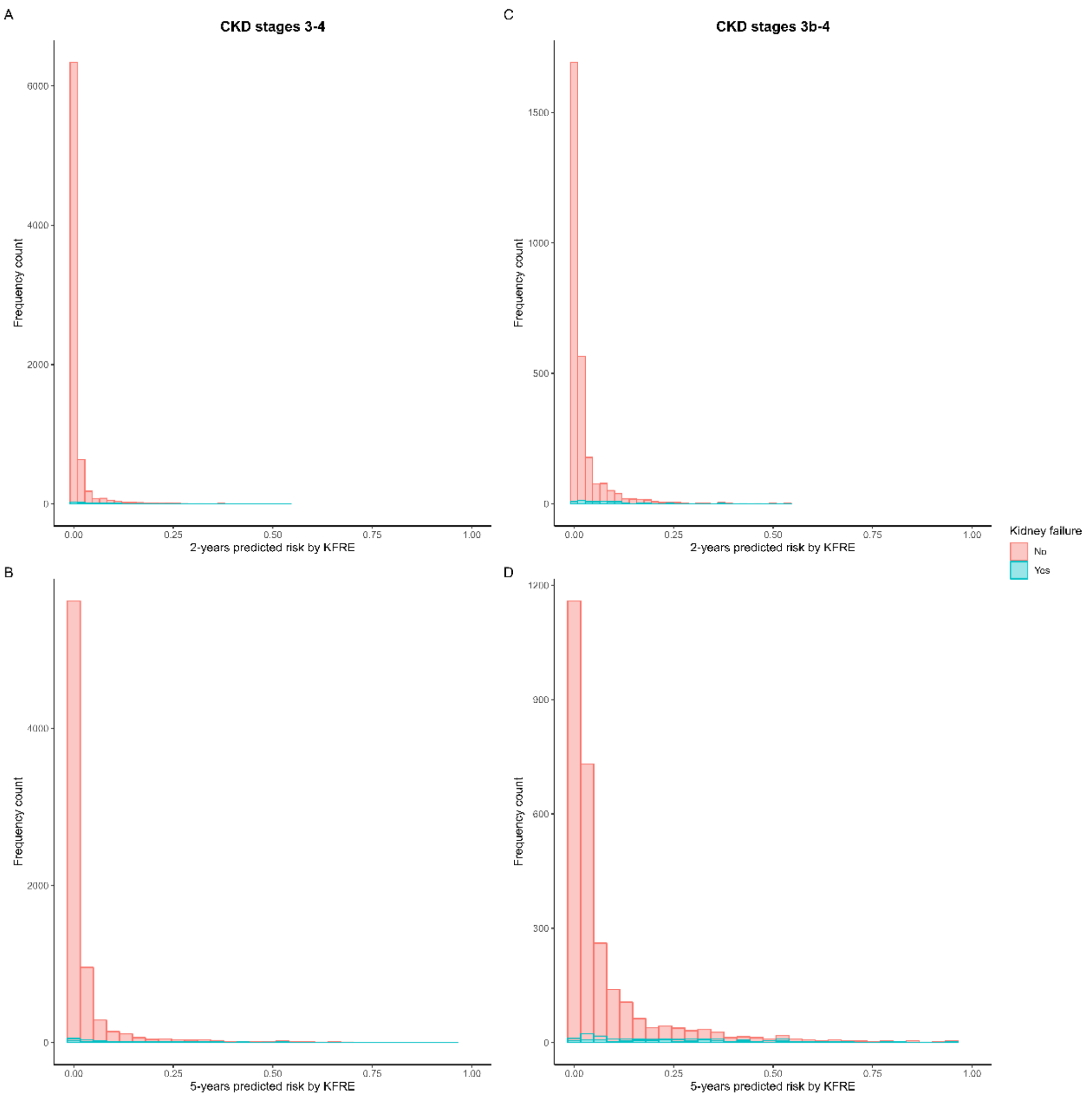

**Figure S1. Distribution of the 2-year and 5-year predicted risk estimated by KFRE equation according to kidney failure outcomes.**

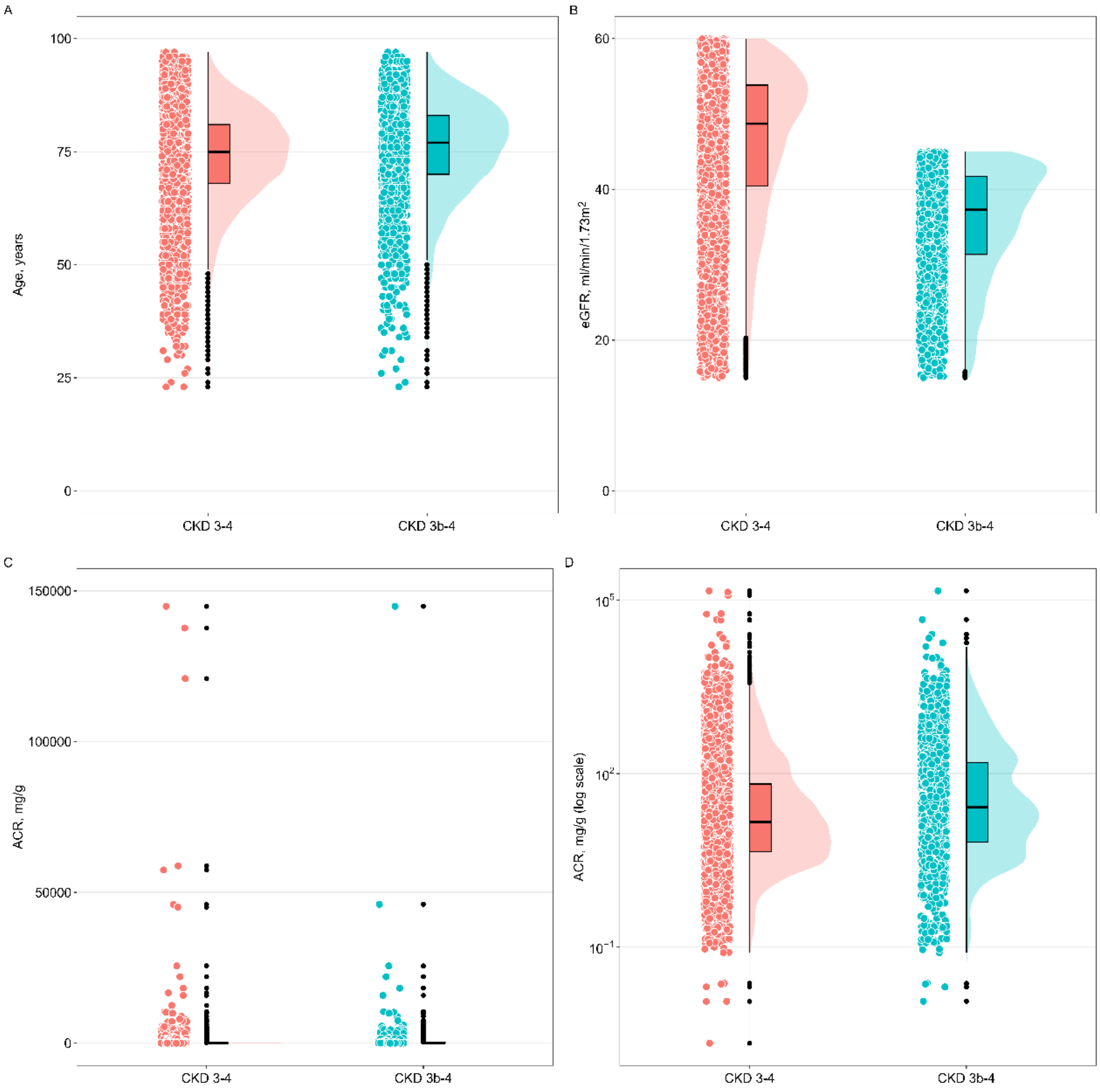

**Figure S2. Distribution of the four KFRE equation variables in CKD 3-4 and CKD 3b-4 patients.**

(A) age in years, (B) estimated glomerular filtration rate (eGFR) according to the CKD-EPI formula, (C) urine albumin to creatinine ratio (ACR) expressed in the original scale, and (D) natural logarithm scale for improved comparison of distributions.

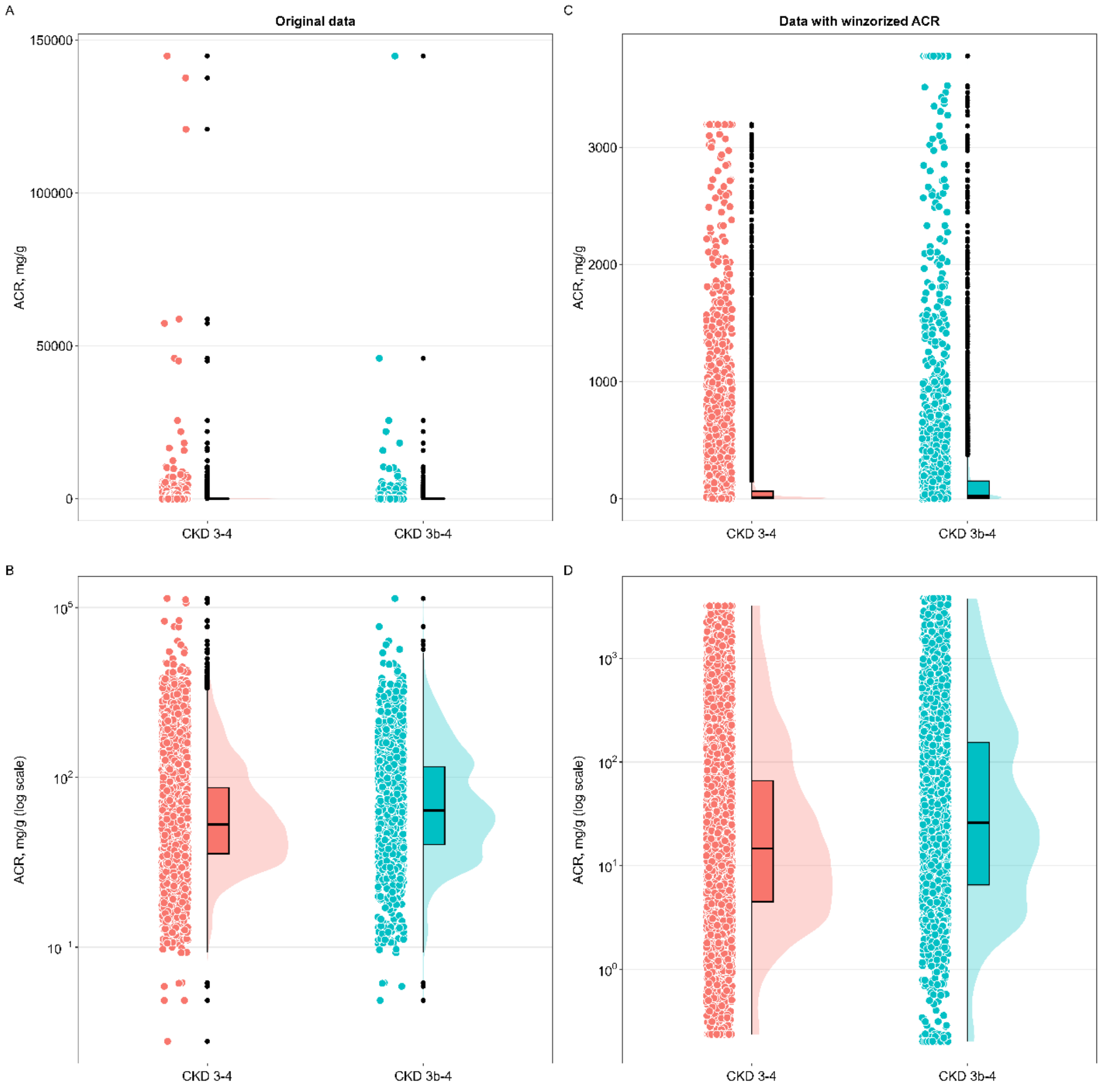

**Figure S3. Distribution of ACR in CKD 3-4 and CKD 3b-4 patients before and after winsorising the 1% and 99% extreme values of ACR.**

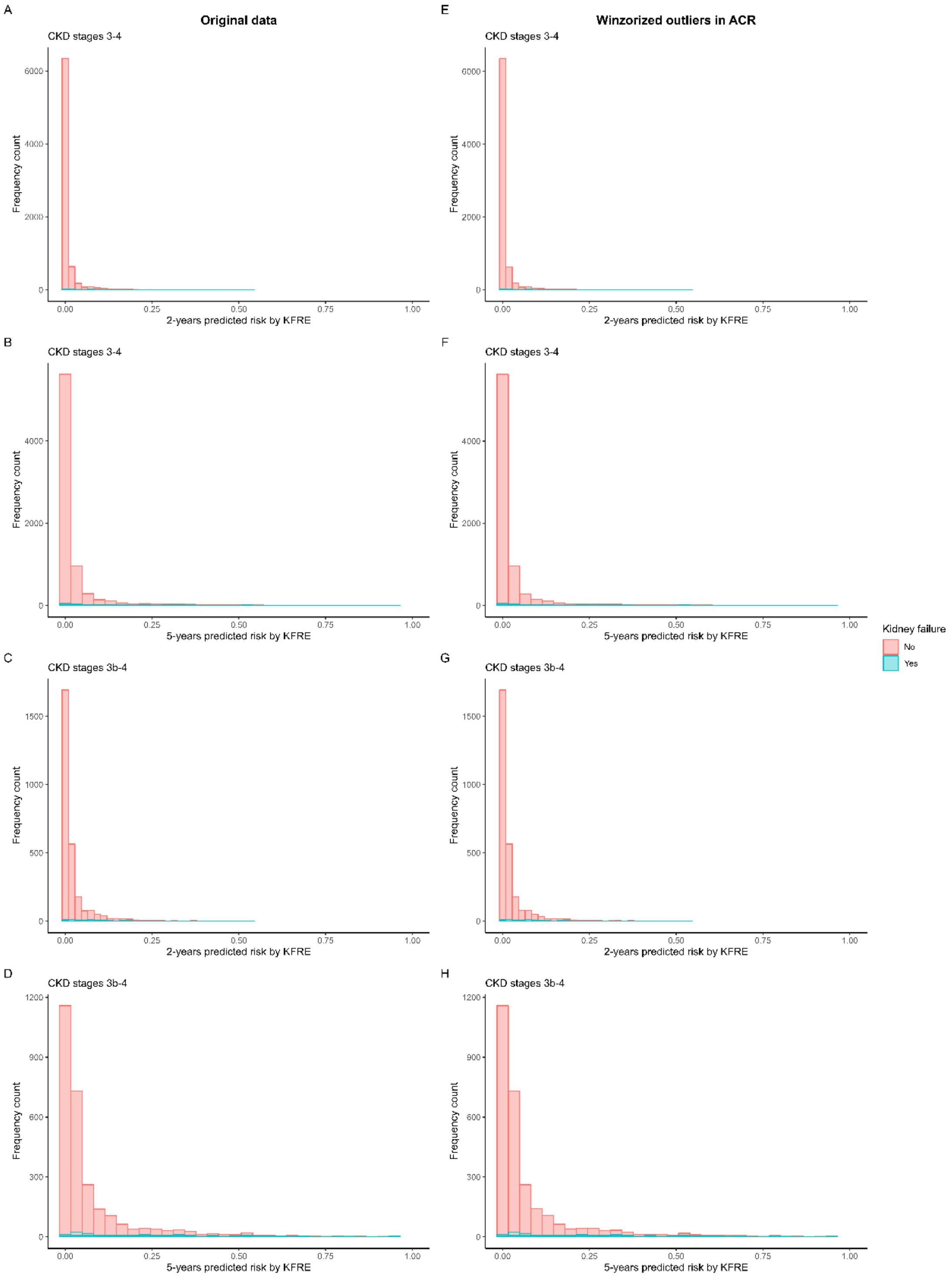

**Figure S4. Distribution of the recalculated 2-year and 5-year predicted risk estimated by the KFRE equation after winsorising the 1% and 99% extreme values of ACR.**

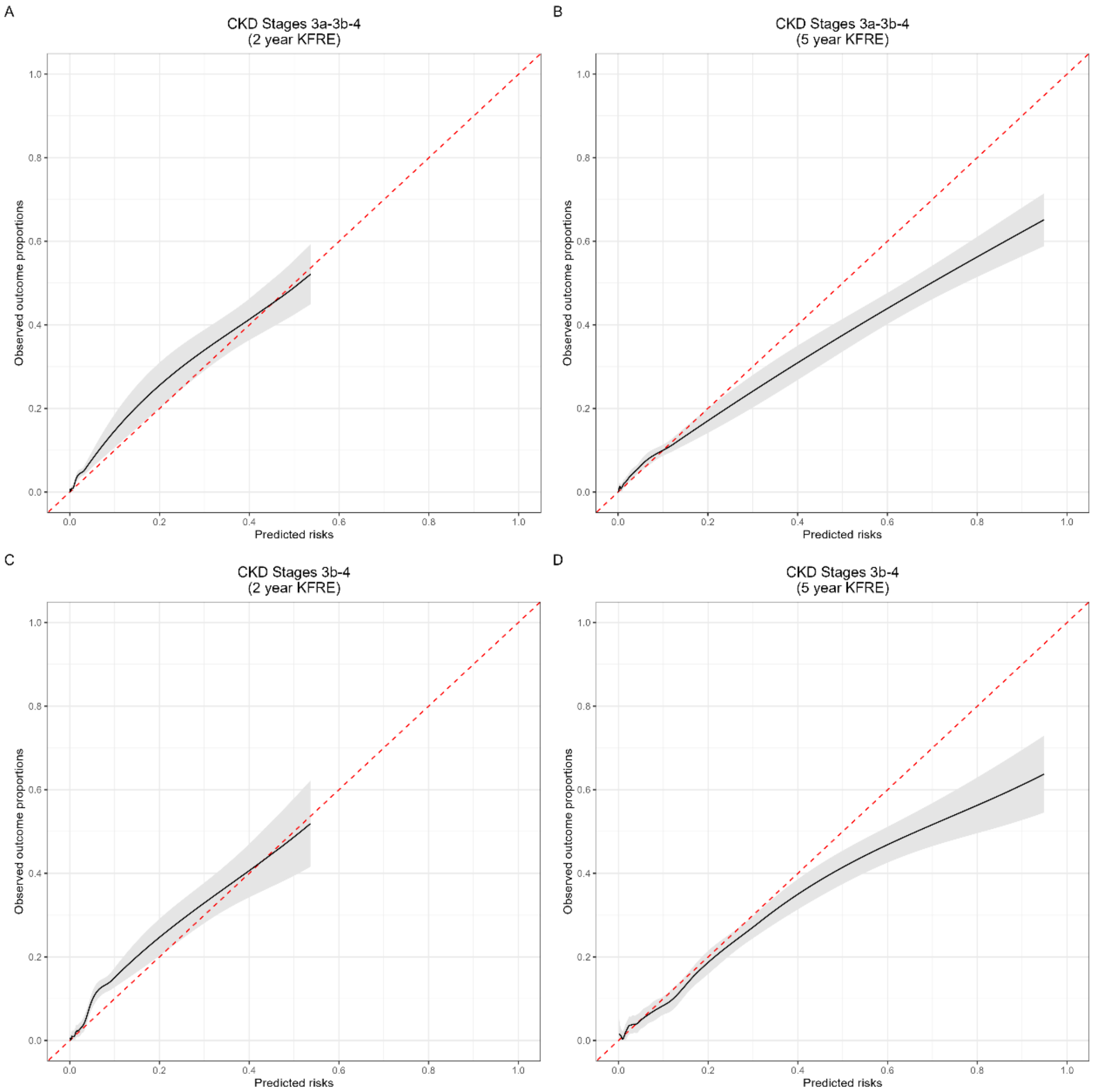

**Figure S5. Calibration curves for each group and prediction horizon following the application of winsorisation to the ACR variable and the subsequent recalculation of predicted risks using the Kidney Failure Risk Equation (KFRE).**

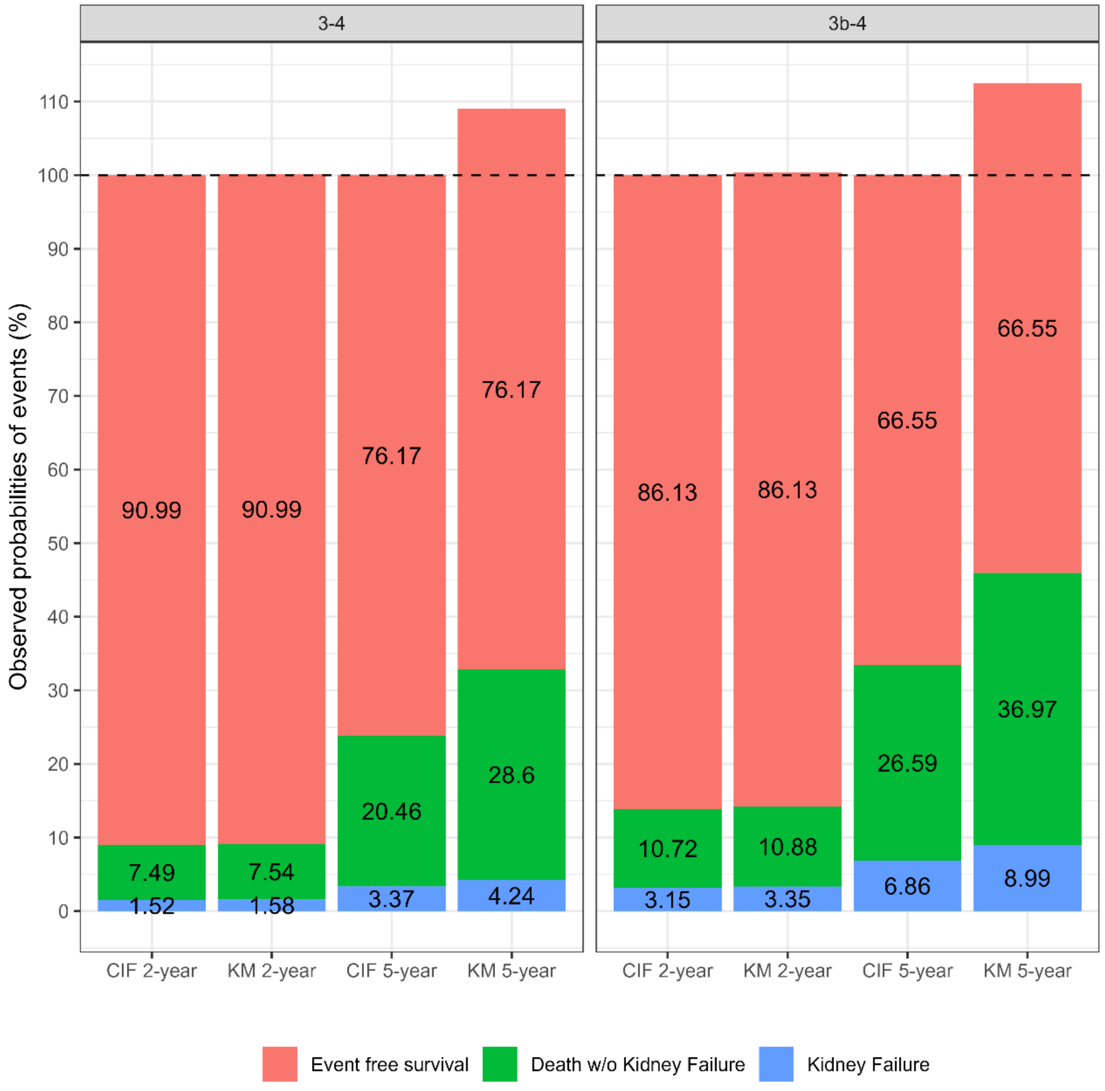

**Figure S6. Differences between Kaplan-Meier (KM) and cumulative incidence functions (CIF) estimates of the observed outcome risks in the presence of competing events, in CKD 3a-4 and CKD 3b-4 patients.**

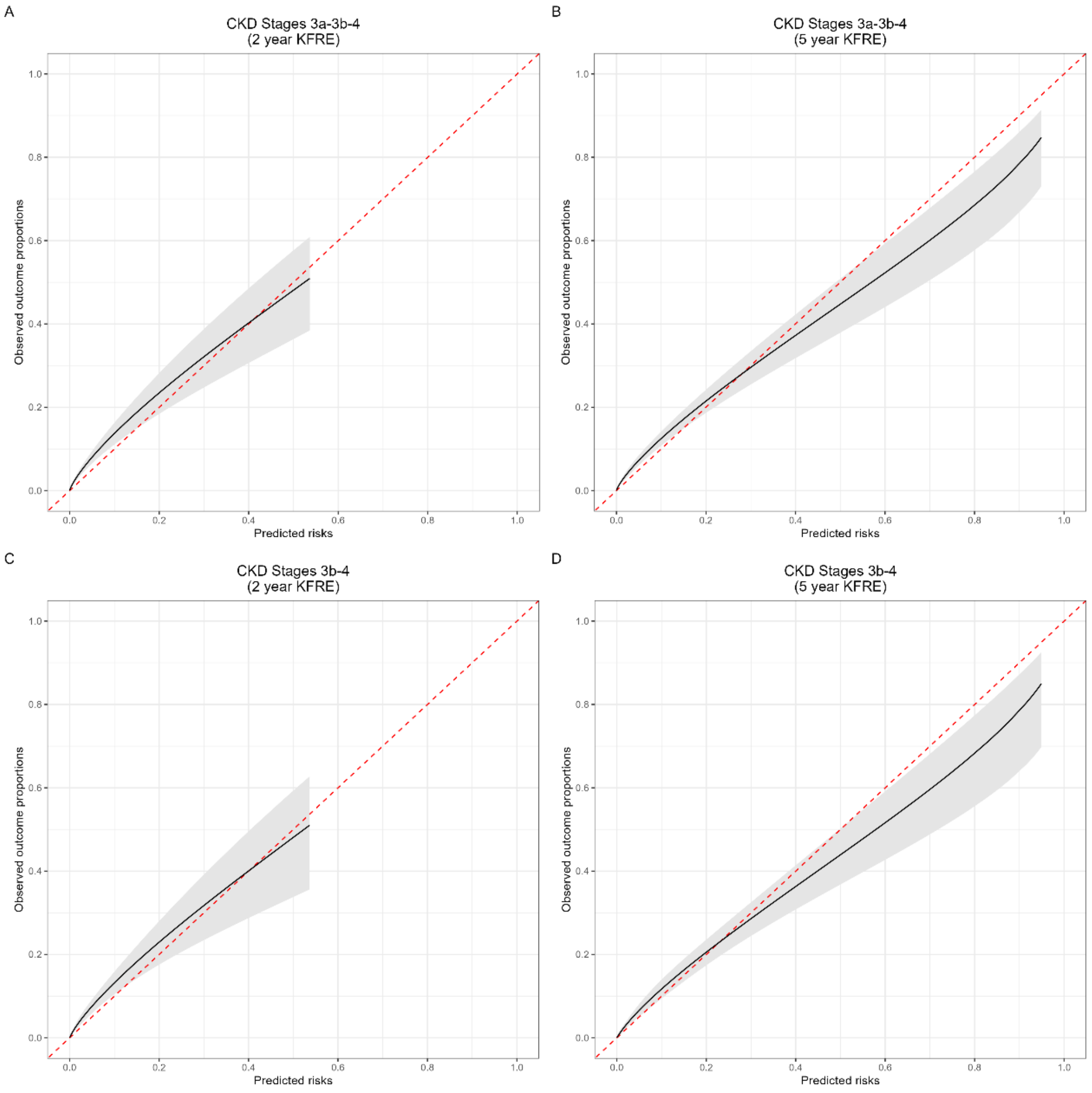

**Figure S7. Calibration curves for each group and prediction horizon, disregarding competing risks.**
